# Maternal Antibodies to Primary CMV Infection Link to Fetal Transmission

**DOI:** 10.64898/2026.09.10.26362772

**Authors:** Chelsea M. Crooks, Richard T. Barfield, Adelaide S. Fuller, Brenna L. Hughes, Geeta K. Swamy, Sergey Ananyev, Libby Mitchell, Megan R. Connors, Krithika P. Karthigeyan, Claire E. Otero, Frances Saccoccio, Joshua Eudailey, Carolyn Weinbaum, Cliburn Chan, Sallie R. Permar

**Author notes:** Corresponding Author, 525 East 68th Street, Box 225, New York, NY 10065.

## Abstract

**BACKGROUND:** Human cytomegalovirus (HCMV) is the leading global infectious cause of birth defects. Despite its prevalence as a cause of life-long disabilities, there are no licensed vaccines to prevent congenital CMV (cCMV), and correlates of protection remain poorly understood. To address this, we identified humoral immune responses to primary HCMV infection during early pregnancy associated with odds of vertical transmission.

**METHODS:** Maternal plasma was collected from a trial that screened >200,000 and enrolled 399 pregnant women serologically diagnosed with primary HCMV infection prior to randomization for hyperimmunoglobulin or placebo therapy (NCT01376778), where 78 transmitted cCMV. Plasma IgG, IgM, and IgA binding to HCMV antigens, neutralization, antibody-dependent cellular phagocytosis, and inhibition of cell-associated viral spread were measured and related to transmission outcome.

**RESULTS:** In a case-control conditional logistic regression, IgM binding to HCMV entry glycoproteins was associated with increased odds of cCMV, identifying potential biomarkers of congenital infection. In contrast, IgG binding to UL16, UL141, prefusion-like gB, gB antigenic domains (AD) 4+5, and gH/gL were associated with decreased odds of cCMV. Surprisingly, IgG binding to the immunodominant gB AD-1 region, pp150, and cell-associated gB were associated with increased odds of cCMV. A LASSO regression model selected IgG responses against UL16 and gB AD-4+5 as associated with decreased odds of transmission, suggesting their potential importance as vaccine targets.

**CONCLUSION:** UL16 and domain specificity of gB-specific IgG responses differentially impact cCMV odds following acute infection, emphasizing the importance of rational antigen selection and gB conformation for HCMV vaccine design to eliminate cCMV.

## Introduction

Human cytomegalovirus (HCMV) is the leading infectious cause of congenital defects and hearing loss in infants infected *in utero*, with approximately 1 in 200 children born with congenital CMV (cCMV) in the United States^1^. Despite high prevalence and life-long morbidity, there are no licensed vaccines for cCMV, partially due to a lack of defined immune correlates of protection against cCMV^2^. The recent failure of Moderna’s phase III trial of a gB and pentameric complex (PC) mRNA vaccine to provide protection against HCMV acquisition, despite induction of robust neutralizing antibodies, underscores the need to identify protective immune responses for rational vaccine design^3^. Risk of vertical HCMV transmission following infection during pregnancy differs between primary (30-40% risk of vertical transmission) and non-primary infection (1-3% risk),^4,5^ suggesting that maternal immunity provides partial protection against cCMV. While work to date has identified potentially protective responses in both vaccinees and pregnant women, the immune mechanisms behind this protection remain poorly defined^6–9^.

To define humoral responses associated with vertical HCMV transmission odds following primary infection, we leveraged plasma samples from 399 pregnant women diagnosed with primary HCMV infection prior to 24 weeks gestation enrolled in a trial of a hyperimmune globulin (HIG) for the prevention of cCMV (NTC01376778)^10,11^, with 78 women vertically transmitting HCMV. Although the trial was stopped for futility, this provided an invaluable opportunity to characterize immune responses in a large cohort of women who did and did not vertically-transmit CMV following primary infection in early pregnancy.

## Methods

### CASE-CONTROL COHORT SELECTION

Plasma was collected at randomization from participants in Clinical Trial NCT01376778^10^ (Table 1, Figure S1, Table S7 in the Supplementary Appendix), which enrolled women with singleton pregnancies and serologically determined primary HCMV infection prior to 24 weeks gestation. In the trial, participants were randomized in a 1:1 ratio to receive monthly infusions of placebo or CMV HIG (Cytogam, CSL Behring). Of the 399 women enrolled, 78 women had infants with cCMV, defined as fetal loss with pathologic evidence of CMV infection or detection in amniotic fluid and/or infant urine or saliva prior to 21 days of life. For this immune marker analysis, cases (women with confirmed transmission) and controls (women without confirmed transmission) were matched^12^ on demographic features that could impact HCMV exposure: treatment group (HIG treatment or placebo), race (White, Black, Other/Mixed/Not reported), ethnicity (Hispanic/Latino or Non-Hispanic/Latino), maternal age (within 10 years), gestational age (within 5 weeks), and parity (nulliparous or multiparous) (Table 1). 71 cases were matched with two controls, while three cases only had one suitably matched control. 22 controls were matched to more than one case. Four cases did not have any suitably matched controls and were excluded.

**Table 1.** Participant Characteristic.

| Characteristic | N | Non transmitter | Transmitter |
| --- | --- | --- | --- |
|  |  | N = 111 (60%)<sup>1</sup> | N = 74 (40%)<sup>1</sup> |
| **Matching Criteria** |  |  |  |
| Treatment Group | 185 |  |  |
| Active |  | 62 (56%) | 42 (57%) |
| Placebo |  | 49 (44%) | 32 (43%) |
| Maternal Age (randomization, years) | 185 | 27.0 (22.0, 32.0) | 26.5 (20.0, 31.0) |
| Maternal Race | 185 |  |  |
| Black/African American |  | 19 (17%) | 21 (28%) |
| Other race/More than one race/Not Reported |  | 15 (14%) | 10 (14%) |
| White |  | 77 (69%) | 43 (58%) |
| Maternal Ethnicity | 185 |  |  |
| Hispanic/Latino |  | 21 (19%) | 13 (18%) |
| Not Hispanic/Latino |  | 90 (81%) | 61 (82%) |
| Gestational Age (randomization, weeks) | 185 | 15.7 (12.9, 18.1) | 16.5 (13.3, 19.6) |
| Parity | 185 |  |  |
| Multiparous |  | 69 (62%) | 42 (57%) |
| Nulliparous |  | 42 (38%) | 32 (43%) |
| **Additional Maternal Demographics** |  |  |  |

**Table 1. Participant Characteristics**
| Characteristic | N | Non transmitter | Transmitter |
| --- | --- | --- | --- |
|  |  | N = 111 (60%)<sup>1</sup> | N = 74 (40%)<sup>1</sup> |
| Days between screening and randomization | 185 | 24 (19, 33) | 23 (18, 29) |
| Risk of Home/Occupational Exposure to CMV | 185 | 86 (77%) | 64 (86%) |
| Maternal BMI (randomization) | 185 | 26 (23, 30) | 26 (22, 29) |
| **Infant Demographics** |  |  |  |
| Gestational Age (delivery, weeks) | 185 | 39.4 (38.6, 40.3) | 38.5 (36.6, 39.3) |
| Pre-term delivery, <37 weeks | 185 | 3 (2.7%) | 21 (28%) |
| Pre-term delivery, <34 weeks | 185 | 1 (0.9%) | 10 (14%) |
| Neonate Head Circumference (delivery, cm) | 176 | 34.20 (33.00, 35.00) | 33.50 (32.50, 34.90) |
| Neonate Weight (delivery, g) | 177 | 3,360 (2,980, 3,635) | 3,098 (2,630, 3,464) |
| Infant Sensorineural Hearing Loss (4 weeks) | 129 | 0 (0%) | 6 (12%) |
| Symptomatic CMV infection | 185 |  |  |
| CMV infection and at least one symptom |  | 0 (0%) | 35 (47%) |
| CMV infection and no symptoms |  | 0 (0%) | 39 (53%) |
| No CMV infection |  | 111 (100%) | 0 (0%) |
| Fetal or neonatal death | 185 | 0 (0%) | 9 (12%) |
| Termination of pregnancy | 185 | 0 (0%) | 5 (6.8%) |

**Table 1. Participant Characteristics**
| Characteristic | N | Non transmitter | Transmitter |
| --- | --- | --- | --- |
|  |  | N = 111 (60%)<sup>1</sup> | N = 74 (40%)<sup>1</sup> |

<sup>1</sup>n (%); Median (Q1, Q3)

### MEASUREMENT OF HUMORAL IMMUNE MARKERS AND QUANTIFICATION OF DNAEMIA

Samples used for analysis were collected at participants’ first infusion, prior to administration of HIG or placebo, which occurred 24.56 ± 8.45 (mean ± SD) days after screening for primary infection. 57 humoral immune markers were chosen to assess both the specificity and function of the maternal plasma antibody responses against HCMV (Table S2)^7,13–15^. DNAemia was measured via Immediate Early 1 (IE1) gene qPCR, as previously reported, on the same plasma samples^11^. Personnel performing assays were blinded to participants’ transmission status. Details about immunological assays and DNAemia detection are in the Supplementary Appendix.

### STATISTICAL ANALYSIS

Prior to conducting this study, a 1:2 match with 78 cases, with an effect size of d=0.4 and an unadjusted p-value significance level of 0.05 was estimated to have 82% power to detect immune markers associated with transmission odds. We performed univariate conditional logistic regression likelihood ratio tests for each immune marker’s association with vertical transmission odds in our case-control cohort^16^. Multiple testing correction was performed by assay type via Benjamini-Hochberg’s False Discovery Rate (FDR), with significance set at a multiple-testing adjusted p-value (q) less than 0.20 and an unadjusted p-value (p) less than 0.05^17^. Spearman correlations were calculated for all immune markers restricted to pairs with non-missing data. Exploratory analysis tested the differences in the distribution for each immune marker between participants with and without DNAemia via Wilcoxon rank-sum test^18^. We also assessed whether, amongst participants with DNAemia, DNAemia magnitude changed transmission odds. We evaluated the markers jointly by performing 100 imputations of data missing due to failure to meet quality control criteria. Imputed datasets were then assessed via a conditional logistic LASSO^19^ with five-fold cross-validation to select the model that minimized mean cross-validation deviance^20,21^. Feature importance was determined by examining the medium effect size observed over each imputation^22^. Sensitivity tests were performed for both univariate analysis and LASSO analysis as described in the Supplementary Appendix.

## Results

### UNIVARIATE ANALYSIS OF IMMUNE MARKERS WITH VERTICAL TRANSMISSION ODDS

We successfully measured immune responses within a dynamic range for 55 of 57 immune markers tested (Figs. S2-S10). Neutralization of the Towne strain in fibroblasts with and without complement was detectable in <20 of 185 participants and thus were excluded from further analysis (Fig. S8).

20 HCMV antigen binding plasma antibody responses were associated with increased (n=13) or decreased (n=7) odds of vertical transmission in our case-control cohort (Figure 1, Table S2). Plasma IgG avidity (p<0.001, q<0.001), but not binding magnitude (p=0.2, q=0.2), to whole virions (AD169r strain) was associated with reduced transmission odds. No IgM responses were associated with lower odds of transmission. However, the magnitude of IgM binding to both full-length entry glycoproteins and antigenic domains (ADs) of gB was associated with increased transmission odds, including gB ectodomain (p<0.001, q<0.001), full-length postfusion gB (p<0.001, q=0.002), PC (p<0.001, q<0.001), gB AD-1 (p<0.001, q=0.002), gB AD-4+5 (p=0.02, q=0.04), and gB AD-4 (p=0.05, q=0.08). IgM binding to two tegument proteins (pp150, p<0.001, q= 0.002; pp71, p=0.003, q=0.007) and to UL16 (p=0.008, q=0.02) was also associated with higher odds of transmission. Plasma IgA binding only to pp65 was associated with increased transmission odds (p=0.002, q=0.03). The magnitude of IgG binding to UL16 (p=0.001, q=0.004), UL141 (p=0.04, q=0.09), prefusion-like gB (p=0.05, q=0.09), gB AD-4+5 (p<0.001, q=0.002), gB AD-5 (p<0.001, q=0.002), and gH/gL (p=0.004, q=0.009) was associated with decreased odds of transmission. Surprisingly, IgG binding to gB AD-1 (p<0.001, q<0.001), pp150 (p<0.001, q<0.001), and cell-associated gB (p=0.005) was associated with increased odds of transmission. Post hoc analysis of UL16- and gB AD-4+5-specific IgG levels showed significant association with reduced transmission odds (p<0.001), while prefusion-like gB-specific IgG was not associated with transmission odds (p=0.11) (Figure S10).

**FIGURE 1.**
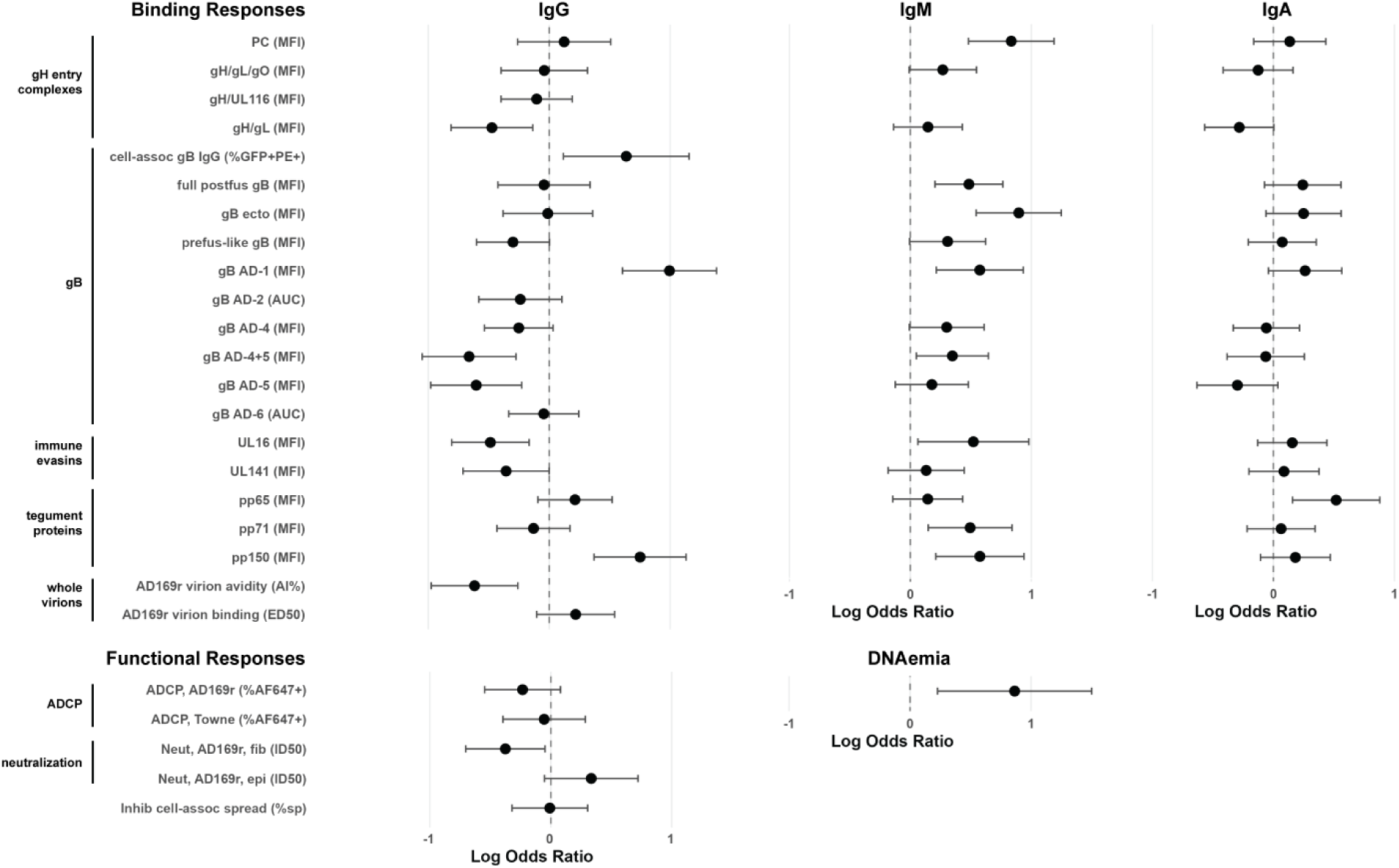
UNIVARIATE ANALYSIS OF PLASMA HCMV-ANTIGEN-SPECIFIC BINDING IMMUNE MARKERS, FUNCTIONAL IMMUNE MARKERS, AND DNAEMIA. Normalized log odds ratio with 95% confidence interval (conditional logistic regression) is plotted for each immune marker. DNAemia log odds ratio is calculated based on the presence or absence of DNAemia in each participant. Positive log odds ratio indicates association with increased odds of transmission; negative log odds ratio indicates association with decreased odds of transmission. Assay units indicated in parentheses. Multiple testing correction was performed with the Benjamini-Hochberg false discovery rate (FDR) for markers of the same assay type and isotype. Data presented from the following assays: binding antibody multiplex assay (BAMA; median fluorescence intensity; MFI), ELISA (area under the curve; AUC), binding to cell-associated gB (%GFP+%PE+ cells); whole virion ELISA binding (ED_50_) and avidity (avidity index; AI%), antibody dependent cellular phagocytosis (ADCP; %AF647+ cells), HCMV neutralization (ID_50_), inhibition of cell-associated viral spread (% spread), presence of HCMV DNAemia (IE1 qPCR).

Only one functional response, neutralization of AD169r on fibroblasts, was associated with decreased transmission odds (p=0.02, q=0.03) (Figure 1, Table S2). While not significant, neutralization of AD169r on epithelial cells had an estimated increase in transmission odds (p=0.06, q=0.06). Antibody-dependent cellular phagocytosis and inhibition of cell-associated viral spread were not associated with transmission odds.

### CORRELATION OF BINDING AND FUNCTIONAL ANTIBODY RESPONSES

Antigen-specific antibody binding and functional responses (neutralization, ADCP, inhibition of cell-associated viral spread) only demonstrated a modest correlation (Figure 2, Table S6). Spearman correlation with functional responses did not exceed ρ<±0.57 for IgG binding levels, ρ<±0.32 for IgM binding, and ρ<±0.23 for IgA binding. IgG responses tended to be positively correlated with functional responses, whereas IgM tended to be negatively correlated with functional responses. IgA binding responses had a mixture of positive and negative correlations.

**FIGURE 2.**
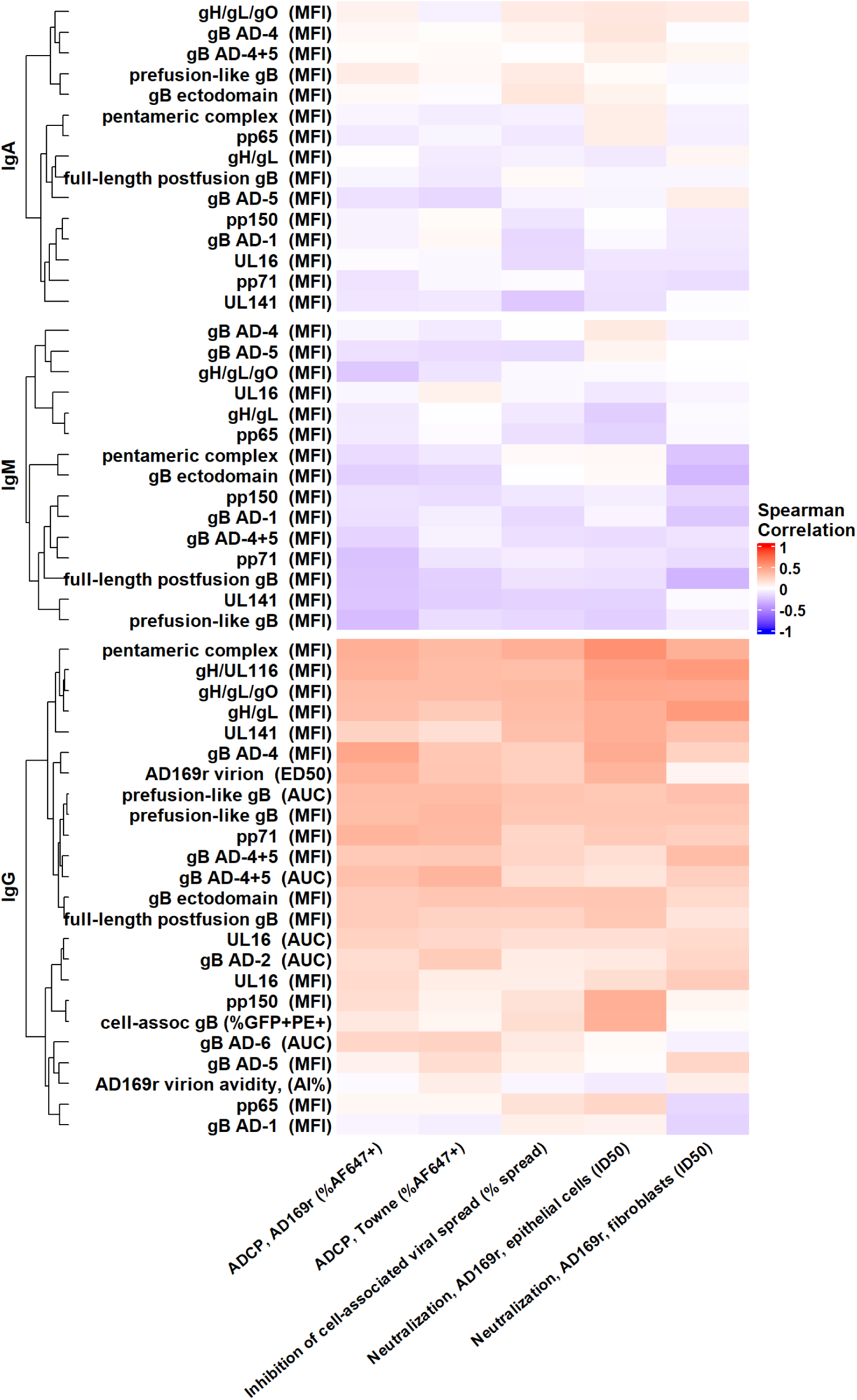
CORRELATION BETWEEN PLASMA HCMV-SPECIFIC BINDING AND FUNCTIONAL ANTIBODY RESPONSES. Spearman correlation was calculated for each plasma antibody binding (left) and functional (bottom) response. Assay units indicated in parentheses. Heatmap and hierarchical clustering generated using the heatmap package in R.

### ASSOCIATION OF DNAEMIA WITH VERTICAL TRANSMISSION ODDS

DNAemia at randomization was quantified for 183 of 185 participants via IE1 gene-specific qPCR (Fig. S11). 81 participants had detectable DNAemia at randomization with a range of 66 to 38,693 copies/mL (median 978 copies/mL). Detectable DNAemia was previously identified as a factor associated with increased odds of transmission in a four-factor model assessing non-invasive predictors of congenital transmission in the trial cohort^11^. In our case-control cohort, detectable DNAemia was associated with increased odds of transmission (p=0.006). However, when testing the impact of DNAemia magnitude on transmission odds in participants with quantifiable DNAemia, there was not a significant association between the magnitude of DNAemia and transmission odds (Wald test, p=0.13, Table S4). Given the association between DNAemia presence and increased transmission odds, all significant univariate analyses were repeated adjusting for the presence of DNAemia, with all remaining significant in this sensitivity analysis (Table S3).

We next assessed differences in immune markers between participants with and without DNAemia. Eleven of 55 immune markers were significantly different between participants with and without DNAemia (Figure 3, Table S5). Nine immune markers were higher in participants with DNAemia; however, plasma UL16-specific IgG (across two assay types) and UL141-specific IgG were higher in participants without DNAemia.

**FIGURE 3.**
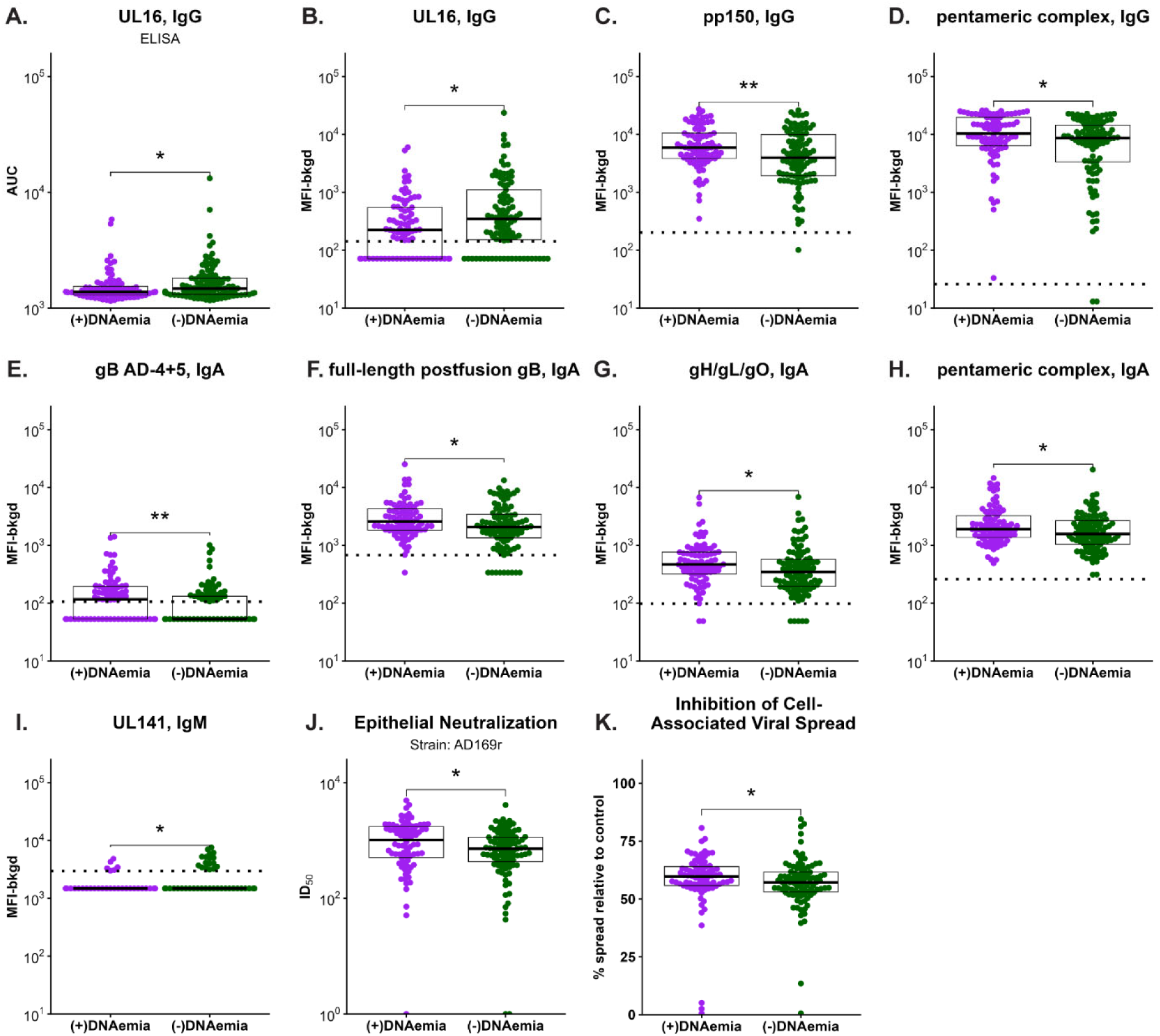
HUMORAL IMMUNE RESPONSES TO ACUTE HCMV INFECTION THAT ARE SIGNIFICANTLY DIFFERENT BETWEEN PARTICIPANTS WITH AND WITHOUT CONCURRENT DNAEMIA. Wilcoxon rank-sum test was used to assess differences in immune markers between participants with and without concurrent DNAemia. The 11 immune markers with p<0.05 are shown here including both binding (**A-I**) and functional (**J, K**) responses. Most immune markers were higher in participants with DNAemia, but IgG binding to UL16 (ELISA data shown in **A**, multiplex binding assay shown in **B**) and IgM binding to UL141 (**I**) were higher in participants without DNAemia. Dashed line indicates positivity threshold for multiplex binding assay (**B-I**). ID_50_ for participants without detectable neutralization were set to ID_50_=0 for analysis and ID_50_=1 for visualization on a logarithmic scale (**J**).

### MULTIVARIABLE MODEL SELECTS FEATURES ASSOCIATED WITH REDUCED TRANSMISSION ODDS

We performed conditional logistic LASSO to jointly assess the immune markers association with transmission. A marker’s importance in reducing the conditional logistic model’s error was determined via the medium effect size observed over all imputations^21^. Ten markers were selected in more than 50 of 100 imputations (Figure 4). To assess the robustness of the features selected by the LASSO model, three sensitivity analyses were performed: all but one of the highly correlated variables (ρ>0.8) were excluded as this correlation may bias the features that were selected (Fig S12); DNAemia was included to determine whether this impacted the importance of immune markers (Fig. S13); and, the model was run only including participants with data that passed quality control for all 55 variables (Fig S14). Across sensitivity analyses, eight immune markers were consistently selected with the same direction, but different magnitude, of the effect size. The markers include both protective (gB AD-4+5, gB AD-5, and UL16-specific IgG) and deleterious (gB ectodomain, UL16, and PC-specific IgM; pp150 and gB AD-1-specific IgG) immune markers. The stability of these eight highly selected variables across the sensitivity analyses underscores their importance in predicting vertical transmission.

**FIGURE 4.**
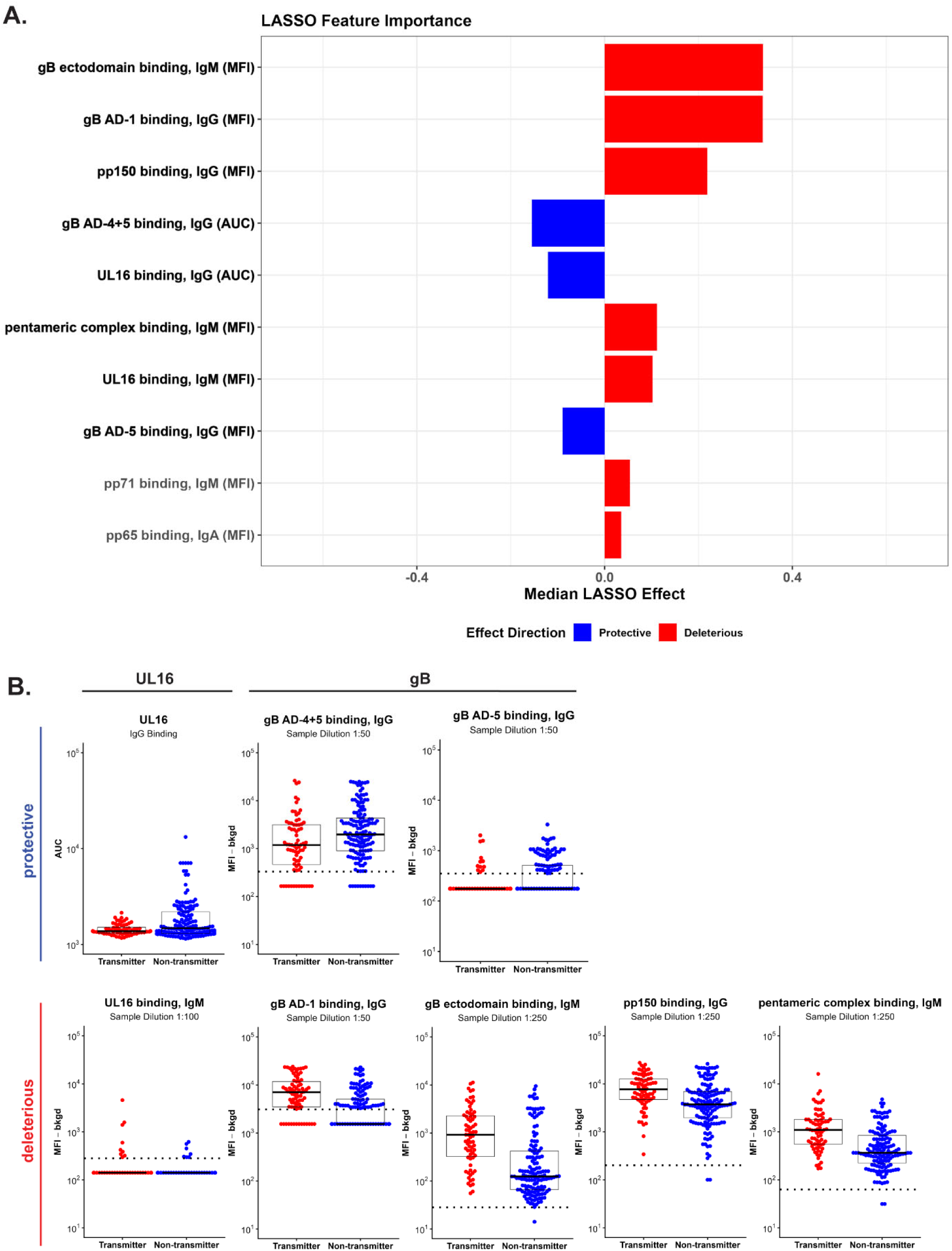
CONDITIONAL LOGISTIC REGRESSION LASSO ANALYSIS OF MATERNAL ANTIBODY RESPONSES ASSOCIATED WITH cCMV TRANSMISSION. Median LASSO effect for immune markers selected in conditional logistic regression LASSO analysis (**A**). Three sensitivity analyses were performed: pruning highly correlated variables (ρ > 0.8), including DNAemia as a variable, and using only participants with data for all 57 immune markers (**Figs. S12-S14**). Eight immune markers with the highest median LASSO effect were stable across all sensitivity analyses. Dot plots for these eight variables (bolded in **A**) are shown in **B**. Dashed line indicates the positivity threshold for a given assay.

## Discussion

Here we report the maternal humoral immune responses associated with vertical transmission odds following primary HCMV infection in early pregnancy. Our matched case-control cohort was well-powered to identify humoral immune responses associated with protection against cCMV. IgG responses to UL16 and gB AD-4+5 were consistently associated with reduced transmission odds, whereas IgG responses against pp150 and gB AD-1 and IgM responses against gB ectodomain, UL16, and PC were associated with increased transmission odds.

It is thought that gB will need to be included in any successful HCMV vaccine^3^. Prior candidates have used gB constructs that include all non-cytosolic antigenic domains of gB identified to date^4,22–26^. Interestingly, in this cohort, responses to specific antigenic domains, but not full-length or ectodomain constructs, were associated with vertical transmission odds. While there is minimal correlation between gB AD-1 and AD-4+5-specific IgG responses, there are very few participants that have both a high gB AD-1 and high AD-4+5-specific IgG response (Fig. S15), indicating an opportunity to build on natural immunity, which is not fully protective against cCMV. The mechanisms underlying the development of a predominantly gB AD-1 or AD-4+5-specific IgG response remain unclear but provide a rationale for gB antigen design.

UL16 is a highly expressed immunoevasin glycoprotein that sequesters NKG2D ligands to evade NK-mediated killing^27^. Antibodies directed at UL16 have been identified as mediating antibody-dependent cellular cytotoxicity (ADCC)^28^ and prior work in maternal-cord blood dyads identified UL16-specific antibodies, as well as engagement of FcɣRIII and ADCC, as associated with lower risk of transmission^8^. Our study confirms in both univariate and LASSO conditional logistic regression that plasma UL16-specific IgG is associated with lower odds of vertical transmission. Of note, anti-UL16 IgG responses were lower magnitude than other HCMV glycoproteins, with only 134 of 185 women generating a response above the positivity threshold. This highlights an opportunity for boosting a potentially protective response through vaccination above levels achieved through infection.

Despite identifying multiple binding antibody responses associated with vertical transmission odds, we only identified one functional response, neutralization of the AD169r strain on fibroblasts, with a significant association; however, this response was not selected in the LASSO model. In fact, neutralizing antibodies have not been associated with protection against HCMV acquisition or cCMV in immune correlate analyses^6,7,29^. Identification of protective functional antibody responses may be hampered by viral strains used in functional assays. HCMV is the most diverse of the herpesviruses and regions of hypervariability exist across the genome including within gB, gO, and gH^30,31^. The use of laboratory strains does not consider the diversity of the participants’ infecting strains. Any HCMV vaccine will need to be effective against diverse HCMV strains and future work should seek to define the required breadth of protective responses.

When viewed collectively, these data suggest humoral immune markers of a more mature immune response against HCMV have lower odds of transmission. IgM binding to entry glycoproteins, IgG binding to pp150 and gB AD-1, and neutralization of epithelial cells are associated with both an early response to infection and with increased odds of transmission in this cohort^9,32^. In contrast, IgG binding to gB AD-4+5, higher avidity IgG to HCMV virions, and neutralization in fibroblasts, all responses generated later in infection^9,33,34^, are associated with lower odds of transmission. These associations may result from variable humoral response development speed after acute infection or differences in the timing of infection relative to gestational age. While participants were enrolled based on serological screening for recent primary infection at <24 weeks gestation and matched on gestational age at sampling (within 5 weeks), the precise timing of infection relative to gestational age and sample collection is unknown. However, transmitting and non-transmitting women had similar binding responses across gestational ages at sampling (7.9-27.9 weeks gestation) suggesting the timing of the infection relative to gestational age may not be a driving factor in these associations (Figs. S16-S18). Presence of DNAemia at the time of sample collection, but not the magnitude, was associated with increased odds of transmission, highlighting the importance of virological control for the reduction of transmission odds. Interestingly, IgM responses to two targets of protective IgG responses, gB AD-4+5 and UL16, were associated with increased odds of transmission. Plasma IgM specific for these antigens may block IgG binding, including IgG that mediate ADCP or ADCC, as has been seen with interfering IgA antibodies in an HIV vaccine trial^35^. However, plasma virus-specific IgM concurrent with IgG was protective in other contexts^36^ and this response may instead be a temporal marker of a more recent infection during a vulnerable period of pregnancy. Additional studies are needed to identify both the viral replication kinetics and immunologic mechanisms underlying these associations.

While this unprecedentedly large cohort provides an invaluable opportunity to study humoral responses against cCMV, due to the study design, there are limitations to our conclusions. These data come from a single timepoint, and conclusions cannot be drawn regarding the durability of immune responses or kinetics of DNAemia. This study used a case-control matched cohort design, which allowed testing more variables across a subset of the study population with strong power to detect responses associated with transmission odds. However, given this design, we are unable to report on risk ratios for each immune marker^37^. Furthermore, while maternal antibody responses clearly play a significant role in protection against cCMV, previous work has highlighted the role of both cellular and innate immunity in protection against HCMV acquisition and cCMV disease^38,39^, which were not assessed in this study. And, while primary HCMV infection carries a higher risk of vertical transmission, many cCMV cases result from non-primary infection due to widespread HCMV seroprevalence. Similar studies of women with non-primary infection are needed to determine whether the responses identified here also reduce transmission odds following non-primary infection.

This study identified rational targets for the design of HCMV vaccines with the goal of preventing congenital infection. While this study was not designed to identify formal correlates of protection, logistical and ethical challenges associated with studying infections in pregnancy mean true correlates of protection may be impossible to define for cCMV. All trials to date of vaccines developed to eliminate cCMV have used virus acquisition as an endpoint, but no candidate has demonstrated >50% efficacy against acquisition of this virus that has evolved for millions of years to evade human immune responses. Thus, designing HCMV vaccines around the prevention of cCMV and re-evaluation of the appropriate clinical trial end points is needed to reach the goal of a licensed HCMV vaccine. This study should inform future work to design vaccines that control viral replication and elicit potentially protective IgG responses against cCMV as identified in this study, including UL16 and gB AD-4+5-specific responses.

## Data Availability

All data and code used in the present study are available upon request to the authors.

## Acknowledgements

Thank you to all the participants in Clinical Trial NCT01376778 and to the Eunice Kennedy Shriver National Institute of Child Health and Human Development, the Maternal-Fetal Medicine Units Network and the CMV Protocol Subcommittee for making data available for this research.

We thank members of the Permar lab that processed participant samples. We would like to thank Richard Stanton for providing UL16 and UL141 proteins. We would like to thank Florian Klein and Matthias Zehner for providing gH/UL116 protein. We would like to thank Jason McLellan for providing prefusion-like gB protein. We would like to thank Robert Kalejta for providing the pp71 plasmid.

This work was supported by NIH NIAID 5R01AI173333 “Identifying and modeling immune correlates of protection against congenital CMV transmission after primary maternal infection” to S.R.P.; Hartwell Foundation Postdoctoral Research Fellowship to C.M.C.; National CMV Foundation PIDS Fellowship Award and Thrasher Research Foundation Early Career Award to F.S. Clincal Trial NCT01376778 was supported by grants (UG1 HD40500, U24 HD36801, UG1 HD53097, UG1 HD40512, UG1 HD40485, UG1 HD34208, UG1 HD40560, UG1 HD27869, UG1 HD27915, UG1 HD68258, UG1 HD68282, UG1 HD40544, UG1 HD40545, UG1 HD68268, UG1 HD34116, UG1 HD87192, and UG1 HD87230) from the *Eunice Kennedy Shriver* National Institute of Child Health and Human Development (NICHD) and the National Center for Advancing Translational Sciences (UL1TR001873 and UL1TR000040). This project was supported in part by the Duke Clinical and Translational Science Institute (CTSI) and by the National Center for Advancing Translational Sciences (NCATS), National Institutes of Health, through Grant 1UM1TR005436. Both Cytogam and AlbuRx were provided by CSL Behring, Inc., free of charge. The company had no involvement in the data management, analysis, or preparation of this manuscript.

The views expressed are those of the author(s) and do not represent the official views of or the official policy of the Duke CTSI, National Institutes of Health, the Department of the Army, the Department of Defense, or the U.S. Government. The investigators have adhered to the policies for the protection of human subjects as prescribed in 45 CFR 46.

## References

1. CDC. Cytomegalovirus (CMV) and Congenital CMV Infection. (https://www.cdc.gov/cytomegalovirus/about/index.html).

2. Plotkin SA. Preventing Infection by Human Cytomegalovirus. J Infect Dis 2020;221(Suppl 1):S123–S127. DOI: 10.1093/infdis/jiz448.

3. Permar SR, Schleiss MR, Plotkin SA. A vaccine against cytomegalovirus: how close are we? J Clin Invest 2025;135(1). DOI: 10.1172/JCI182317.

4. Boppana SB, van Boven M, Britt WJ, et al. Vaccine value profile for cytomegalovirus. Vaccine 2023;41 Suppl 2:S53–S75. DOI: 10.1016/j.vaccine.2023.06.020.

5. Rawlinson WD, Boppana SB, Fowler KB, et al. Congenital cytomegalovirus infection in pregnancy and the neonate: consensus recommendations for prevention, diagnosis, and therapy. Lancet Infect Dis 2017;17(6):e177–e188. DOI: 10.1016/S1473-3099(17)30143-3.

6. Jenks JA, Nelson CS, Roark HK, et al. Antibody binding to native cytomegalovirus glycoprotein B predicts efficacy of the gB/MF59 vaccine in humans. Sci Transl Med 2020;12(568):eabb3611. DOI: 10.1126/scitranslmed.abb3611.

7. Semmes EC, Miller IG, Wimberly CE, et al. Maternal Fc-mediated non-neutralizing antibody responses correlate with protection against congenital human cytomegalovirus infection. J Clin Invest 2022;132(16). DOI: 10.1172/JCI156827.

8. Semmes EC, Miller IG, Rodgers N, et al. ADCC-activating antibodies correlate with decreased risk of congenital human cytomegalovirus transmission. JCI Insight 2023;8(13). DOI: 10.1172/jci.insight.167768.

9. Lilleri D, Gerna G, Furione M, Zavattoni M, Spinillo A. Neutralizing and ELISA IgG antibodies to human cytomegalovirus glycoprotein complexes may help date the onset of primary infection in pregnancy. J Clin Virol 2016;81:16–24. DOI: 10.1016/j.jcv.2016.05.007.

10. Hughes BL, Clifton RG, Rouse DJ, et al. A Trial of Hyperimmune Globulin to Prevent Congenital Cytomegalovirus Infection. N Engl J Med 2021;385(5):436–444. DOI: 10.1056/NEJMoa1913569.

11. Rouse DJ, Fette LM, Hughes BL, et al. Noninvasive Prediction of Congenital Cytomegalovirus Infection After Maternal Primary Infection. Obstet Gynecol 2022;139(3):400–406. DOI: 10.1097/AOG.0000000000004691.

12. Sekhon JS. Multivariate and Propensity Score Matching Software with Automated Balance Optimization: The Matching Package for R. J Stat Softw 2011;42(7):1–52. (In English) (<Go to ISI>://WOS:000292097200001).

13. Karthigeyan KP, Connors M, Binuya CR, et al. A human cytomegalovirus prefusion-like glycoprotein B subunit vaccine elicits humoral immunity similar to that of postfusion gB in mice. J Virol 2025:e0217824. DOI: 10.1128/jvi.02178-24.

14. Bialas KM, Westreich D, Cisneros de la Rosa E, et al. Maternal Antibody Responses and Nonprimary Congenital Cytomegalovirus Infection of HIV-1-Exposed Infants. J Infect Dis 2016;214(12):1916–1923. DOI: 10.1093/infdis/jiw487.

15. Hu X, Karthigeyan KP, Herbek S, et al. Human Cytomegalovirus mRNA-1647 Vaccine Candidate Elicits Potent and Broad Neutralization and Higher Antibody-Dependent Cellular Cytotoxicity Responses Than the gB/MF59 Vaccine. J Infect Dis 2024;230(2):455–466. DOI: 10.1093/infdis/jiad593.

16. Therneau T. A package for survival analysis in R. (https://cran.r-project.org/web/packages/survival/index.html).

17. Benjamini Y, Hochberg Y. Controlling the False Discovery Rate - a Practical and Powerful Approach to Multiple Testing. J Roy Stat Soc B 1995;57(1):289–300. (In English). DOI: DOI 10.1111/j.2517-6161.1995.tb02031.x.

18. Hothon T, Winell H, Hornik K, van de Wiel MA, Zeileis A. Conditional Inference Procedures in a Permutation Test Framework.

19. Reid S, Tibshirani R. Regularization Paths for Conditional Logistic Regression: The clogitL1 Package. J Stat Softw 2014;58(12):1–23. (In English) (<Go to ISI>://WOS:000341642900001).

20. van Buuren S, Groothuis-Oudshoorn K. mice: Multivariate Imputation by Chained Equations in R. J Stat Softw 2011;45(3):1–67. (In English) (<Go to ISI>://WOS:000298032500001).

21. Peterson RA. A Simple Aggregation Rule for Penalized Regression Coefficients after Multiple Imputation. Journal of Data Science 2021:1–14. DOI: 10.6339/21-jds995.

22. Pötzsch S, Spindler N, Wiegers A-K, et al. B Cell Repertoire Analysis Identifies New Antigenic Domains on Glycoprotein B of Human Cytomegalovirus which Are Target of Neutralizing Antibodies. PLoS Pathogens 2011;7(8):e1002172. DOI: 10.1371/journal.ppat.1002172.

23. Gomes AC, Baraniak IA, Lankina A, et al. The cytomegalovirus gB/MF59 vaccine candidate induces antibodies against an antigenic domain controlling cell-to-cell spread. Nat Commun 2023;14(1):1041. DOI: 10.1038/s41467-023-36683-x.

24. Meyer H, Sundqvist VA, Pereira L, Mach M. Glycoprotein gp116 of human cytomegalovirus contains epitopes for strain-common and strain-specific antibodies. J Gen Virol 1992;73 (Pt 9):2375–83. DOI: 10.1099/0022-1317-73-9-2375.

25. Wagner B, Kropff B, Kalbacher H, et al. A continuous sequence of more than 70 amino acids is essential for antibody binding to the dominant antigenic site of glycoprotein gp58 of human cytomegalovirus. J Virol 1992;66(9):5290–7. DOI: 10.1128/JVI.66.9.5290-5297.1992.

26. Kniess N, Mach M, Fay J, Britt WJ. Distribution of linear antigenic sites on glycoprotein gp55 of human cytomegalovirus. Journal of Virology 1991;65(1):138–146. DOI: 10.1128/jvi.65.1.138-146.1991.

27. Dunn C, Chalupny NJ, Sutherland CL, et al. Human cytomegalovirus glycoprotein UL16 causes intracellular sequestration of NKG2D ligands, protecting against natural killer cell cytotoxicity. J Exp Med 2003;197(11):1427–39. DOI: 10.1084/jem.20022059.

28. Vlahava VM, Murrell I, Zhuang L, et al. Monoclonal antibodies targeting nonstructural viral antigens can activate ADCC against human cytomegalovirus. J Clin Invest 2021;131(4). DOI: 10.1172/JCI139296.

29. Baraniak I, Kropff B, Ambrose L, et al. Protection from cytomegalovirus viremia following glycoprotein B vaccination is not dependent on neutralizing antibodies. Proc Natl Acad Sci U S A 2018;115(24):6273–6278. DOI: 10.1073/pnas.1800224115.

30. Charles OJ, Venturini C, Gantt S, et al. Genomic and geographical structure of human cytomegalovirus. Proc Natl Acad Sci U S A 2023;120(30):e2221797120. DOI: 10.1073/pnas.2221797120.

31. Lassalle F, Depledge DP, Reeves MB, et al. Islands of linkage in an ocean of pervasive recombination reveals two-speed evolution of human cytomegalovirus genomes. Virus Evol 2016;2(1):vew017. DOI: 10.1093/ve/vew017.

32. Schoppel K, Kropff B, Schmidt C, Vornhagen R, Mach M. The humoral immune response against human cytomegalovirus is characterized by a delayed synthesis of glycoprotein-specific antibodies. J Infect Dis 1997;175(3):533–44. DOI: 10.1093/infdis/175.3.533.

33. Kagan KO, Bissinger AL, Roth O, et al. Predictive value of gB2 antibodies for maternal-fetal transmission after primary cytomegalovirus infection treated with valacyclovir. Ultrasound Obstet Gynecol 2026;67(4):455–460. DOI: 10.1002/uog.70206.

34. Chatzakis C, Ville Y, Makrydimas G, Dinas K, Zavlanos A, Sotiriadis A. Timing of primary maternal cytomegalovirus infection and rates of vertical transmission and fetal consequences. Am J Obstet Gynecol 2020;223(6):870–883 e11. DOI: 10.1016/j.ajog.2020.05.038.

35. Haynes BF, Gilbert PB, McElrath MJ, et al. Immune-correlates analysis of an HIV-1 vaccine efficacy trial. N Engl J Med 2012;366(14):1275–86. DOI: 10.1056/NEJMoa1113425.

36. Ruggiero A, Piubelli C, Calciano L, et al. SARS-CoV-2 vaccination elicits unconventional IgM specific responses in naive and previously COVID-19-infected individuals. EBioMedicine 2022;77:103888. DOI: 10.1016/j.ebiom.2022.103888.

37. Kim HY. Statistical notes for clinical researchers: Risk difference, risk ratio, and odds ratio. Restor Dent Endod 2017;42(1):72–76. DOI: 10.5395/rde.2017.42.1.72.

38. Manuel TD, Mostrom MJ, Crooks CM, et al. A rhesus macaque model of congenital cytomegalovirus infection reveals a spectrum of vertical transmission outcomes. Commun Biol 2025;8(1):1647. DOI: 10.1038/s42003-025-09033-4.

39. Bialas KM, Tanaka T, Tran D, et al. Maternal CD4+ T cells protect against severe congenital cytomegalovirus disease in a novel nonhuman primate model of placental cytomegalovirus transmission. Proceedings of the National Academy of Sciences 2015;112(44):13645–13650. DOI: 10.1073/pnas.1511526112.

