## Supplementary Appendix for "Maternal Antibodies to Primary CMV Infection Link to Fetal Transmission"

### Supplementary Appendix Table of Contents

**LIST OF INVESTIGATORS**

Chelsea M. Crooks, PhD, Department of Pediatrics, Weill Cornell Medicine, New York, NY

Richard T. Barfield, PhD, Department of Biostatistics and Bioinformatics, Duke University School of Medicine, Durham, NC

Adelaide S. Fuller, BS, Department of Pediatrics, Weill Cornell Medicine, New York, NY

Brenna L. Hughes, MD, Department of Obstetrics and Gynecology, Duke University, Durham, NC

Geeta K. Swamy, MD, Department of Obstetrics and Gynecology, Duke University, Durham, NC

Sergey Ananyev, BS, Department of Pediatrics, Weill Cornell Medicine, New York, NY

Libby Mitchell, BS, Department of Pediatrics, Weill Cornell Medicine, New York, NY

Megan R. Connors, BS, Department of Pediatrics, Weill Cornell Medicine, New York, NY

Krithika P. Karthigeyan, PhD, Department of Pediatrics, Weill Cornell Medicine, New York, NY

Claire E. Otero, PhD, Department of Pediatrics, Weill Cornell Medicine, New York, NY

Frances Saccoccio, MD, Children’s Hospital of Richmond at VCU, Richmond, VA

Joshua Eudailey, BS, Department of Pediatrics, Weill Cornell Medicine, New York, NY

Carolyn Weinbaum, BS, Department of Pediatrics, Weill Cornell Medicine, New York, NY

Cliburn Chan, PhD, Department of Biostatistics and Bioinformatics, Duke University School of Medicine, Durham, NC

Sallie R. Permar, MD, PhD, Department of Pediatrics, Weill Cornell Medicine, New York, NY

### SUPPLEMENTARY METHODS

***Trial Sites and Study Personnel For A Randomized Trial to Prevent Congenital Cytomegalovirus (NCT01376778)***

Each clinical center consisted of one or more additional performance sites as listed below. Also listed are members of the Eunice Kennedy Shriver National Institute of Child Health and Human Development Maternal-Fetal Medicine Units Network who contributed to the research.

Clinical Centers:

Brown University, Providence, RI – B. Hughes, D. Rouse, D. Allard, E. Werner, J. Rousseau, L. Beati, J. Milano, J. Lee

University of Texas Medical Branch, Galveston, TX – G. Saade, A. Salazar, L. Pacheco, J. Patel, D. Carlson, K. Smith, A. Nounes, J. DeVolder

Northwestern University, Chicago, IL – G. Mallett, W. Grobman, A. Peaceman

NorthShore University Health System, Evanston, IL – M. Dinsmoor, K. Paycheck

Columbia University, New York, NY – C. Gyamfi-Bannerman, S. Bousleiman, R. Wapner V. Carmona, M. Talucci

Christiana Care, Wilmington, DE – M. Hoffman, A. Vanneman

St. Peter's University Hospital, New Brunswick, NJ – K. Palomares, C. Perez

Drexel University, Philadelphia, PA – L. Plante, C. Tocci,

New York Presbyterian Queens, Flushing, NY – D. Skupski, R. Chan-Akeley

Lehigh Valley Hospital, Allentown, PA

University of Utah Health Sciences Center, Salt Lake City, UT – M. Varner, K. Hill, A. Sowles

LDS Hospital, Salt Lake City, UT – C. Meadows

McKay-Dee Hospital, Ogden, UT – S. Dellerman

Intermountain Med Center, Salt Lake City, UT – L. Hansen, S. Esplin

University of North Carolina at Chapel Hill, Chapel Hill, NC – W. Goodnight, K. Clark, J. Thorp, S. Timlin

WakeMed Health & Hospitals, Raleigh, NC – C. Beamon, H. Byers

Prisma Health, Greenville, SC – K. Eichelberger, A. Moore

University of Alabama at Birmingham, Birmingham, AL – A. Tita, S. Harris, R. Pass, L. Harper, J. Biggio, M. Parks, J. Sheppard

The Ohio State University, Columbus, OH– M. Costantine, A. Bartholomew, M. Landon, J. Iams, C. Shellhaas, K. Markham, B. Rink, C. Buhimschi, F. Johnson, L. Webb

Wright State University Miami Valley Hospital, Dayton, OH – D. McKenna, K. Fennig, K. Snow

Duke University, Durham, NC – G. Swamy, T. Bishop, J. Ferrara

University of Colorado School of Medicine, Anschutz Medical Campus, Aurora, CO – R. Gibbs, K. Hale, K.D. Heyborne, J. Phipers

MetroHealth Medical Center-Case Western Reserve University, Cleveland, OH – E. Chien, W. Dalton

University Hospitals, Cleveland, OH – D. Hackney, A. Mayle

UT Health- University of Texas Health Science Center at Houston, Children's Memorial Hermann Hospital, Houston, TX – S. Chauhan, F. Ortiz, B. Sibai

Stanford University, Stanford, CA – Y. El-Sayed, C. Willson, N. Aziz, D. Lyell, A. Girsen, K. Sherwi

University of Texas Southwestern Medical Center, Dallas, TX – B. Casey, L. Moseley, J. Price, T. Thomas, L. Fay-Randall, A. Sias, M. Garcia

University of Pennsylvania, Philadelphia, PA – S. Parry, J. Craig

University of Pittsburgh, Pittsburgh, PA – H. Simhan, M. Bickus, F. Facco, M. Birsic

Madigan Army Medical Center, Joint Base Lewis-McChord, Tacoma, WA – P. Napolitano, L. Imbruglio, E. Hemman, J. Pates, L. Foglia

Data Coordinating Center: The George Washington University Biostatistics Center, Washington, DC – E. Thom, R. Clifton, C. MacPherson, L. Fete, L. Mele, V. L. Flowers-Fanomezantsoa, T. Boekhoudt

Eunice Kennedy Shriver National Institute of Child Health and Human Development, Bethesda, MD – U. Reddy, C. Spong, S. Pagliaro

MFMU Network Steering Committee Chair (Washington University, Saint Louis, MO) – G. A. Macones, MD, MSCE

#### *Sample processing*

Whole blood was collected from trial participants[1] in a vacutainer tube with EDTA. Samples were shipped on ice and received for processing the day after collection. Whole blood was spun at 2000 rpm for 15 minutes. The plasma layer was removed and aliquoted for storage at -80°C. Plasma samples were heat inactivated at 56°C for 30 minutes and cooled to 4°C prior to use in all immunological assays.

#### *HCMV Recombinant Proteins*

Details about recombinant proteins used in assays can be found in Table S1. For proteins produced in house, protein sequences were codon optimized for mammalian cells, synthesized (Genscript), and then cloned into pcDNA3.1(+) mammalian expression vector (Invitrogen). Plasmids were transfected into suspension cells using a polyethylenimine transfection reagent (Sigma-Aldrich). The supernatant was harvested 2–5 days later and purified using Ni2+-NTA resin (Thermo Fisher Scientific). Purity and identity were confirmed by western blot using HCMV hyperimmune globulin (Cytogam) and/or antigen-specific monoclonal antibodies.

#### *Cell Lines*

FreeStyle™ 293-F Cells (Gibco) were purchased from Thermo Fisher. All other cell lines were obtained from the American Type Culture Collection. Human epithelial kidney (HEK293T) cells used in protein-transfected cell assays were maintained in DMEM containing 10% FBS, 25 mM HEPES buffer, 50 U/mL penicillin, and 50 mcg/mL streptomycin. Human retinal pigment epithelial (ARPE-19) cells used in virus production and neutralization assays were maintained in Dulbecco's modified Eagle medium-12 (DMEM-F12) supplemented with 10% fetal bovine serum (FBS), 50 U/mL penicillin, and 50mcg/mL streptomycin. Human foreskin fibroblast cells (HFF-1) use in virus production and neutralization and cell-associated viral spread assays were maintained in DMEM supplemented with 10% FBS, 25 mM HEPES buffer, 50 U/mL penicillin, 50 mcg/mL streptomycin and gentamicin, and 25 mM L-Glutamine. Human monocyte (THP-1) cells used in antibody-dependent cellular phagocytosis assays were maintained in RPMI-1640 medium containing 10% FBS. All cell lines were maintained for a maximum of 25 passages.

#### *Viruses*

Viruses and bacterial artificial chromosomes (BACs) were a gift from Professor Tom Shenk. Towne virus was propagated on HFF-1 cells in T-175 culture flasks. AD169 revertant virus containing the UL131–UL128 ORF from HCMV TR and expressing GFP (AD169r-BAC-GFP) was propagated from BAC transfection of HFF-1 cells using Lipofectamine-3000 (Thermo Fisher Scientific) to produce seed stocks. Working stocks were propagated from seed stock infection of ARPE-19 cells in T-175 flasks. Ts15nR (epithelial-tropic Towne-BAC-GFP with intact pentamer) was propagated in ARPE-19 cells. The supernatant containing cell-free virus was collected when 90% of cells showed cytopathic effects (~2 weeks) and cleared of cell debris by low-speed centrifugation before ultracentrifugation through a 20% sucrose cushion.

#### *Binding Antibody Multiplex Assay (BAMA)*

Binding Antibody Multiplex Assay (BAMA) was performed as described previously[2-7]. MagPlex magnetic carboxylated fluorescent beads (Luminex) were covalently coupled to purified HCMV antigens. The coupled beads were incubated with maternal plasma in diluent consisting of 1% skim milk, 5% normal goat serum, and 0.05% tween in 1X Dulbecco's phosphate buffered saline. Plasma was diluted 1:1000 (full-length post-fusion gB, gB ectodomain, gH/gL/gO, pentameric complex), 1:250 (prefusion-like gB, pp71, pp150, gB AD-4), or 1:50 (UL141, UL16, pp65, gH/gL, gB AD-5, gB AD-4+5, gB AD-1) for IgG assays; 1:250 (full-length post-fusion gB, gB ectodomain, gH/gL/gO, pentameric complex) or 1:100 (pre-fusion gB, pp71, pp150, gB AD-4, UL141, UL16, pp65, gH/gL, gB AD-5, gB AD-4+5, gB AD-1) for IgM assays; and 1:25 for all antigens for IgA assays. Antigen-specific antibody binding was detected with phycoerythrin-conjugated goat anti-human antibody specific for IgG, IgM, or IgA (2 ug/mL, Southern Biotech).  Samples were read on the Bio-Plex 200 system (BioRad) and results are reported as median fluorescence intensity (MFI). A diluent-only sample was included on each plate to determine bead-specific background binding and reported MFIs are background subtracted. Any plate with a diluent-only well over 125 (IgA, IgM) or 400 (IgG) was repeated.

A serial dilution of HCMV hyperimmune globulin (Cytogam, CSL Behring) and a point dilution of palivizumab (Synagis) were included on each plate as positive and negative controls, respectively, and as an assessment of inter-plate variability. All samples were run in duplicate, and results are reported as the average of both replicates. Any sample with a blank bead background subtracted MFI greater than 1000, an antigen with a coefficient of variation (CV) over 25%, or bead count below 100 was repeated one time. Samples that did not meet QC criteria on either biological replicate were excluded. A positivity threshold was calculated for each antigen as the average + 2 standard deviations (SD) of 13 HCMV seronegative plasma samples. Samples falling below the positivity threshold are set to half of the positivity threshold.

#### *IgG Binding to HCMV Antigens*

Binding to biotinylated linear peptides gB AD-2 and gB AD-6, prefusion-like gB, UL16, and gB AD-4+5 was measured by 384-well plate-based ELISA and reported as the area under the curve as described previously[7]. Briefly, 384 well ELISA plates were coated with 150 ng/well of each peptide in coating buffer (0.1 M NaOH pH 9.6) overnight at 4°C. Plates were blocked with assay diluent (1× PBS [pH 7.4] containing 4% whey, 15% normal goat serum, and 0.5% Tween 20) for 1 hour at room temperature. Three-fold, eight-point serial dilutions for each sample in assay diluent were run in duplicate with a starting dilution of 1:10. Binding was detected using peroxidase-conjugated goat anti-human IgG Fc secondary (Jackson ImmunoResearch). Plates were developed using SureBlue Substrate and $OD_{450}$ was read on the BioTek Synergy LX plate reader. Area under the curve was calculated using the AUC function from the DescTools package in R.

A serial dilution of HCMV hyperimmune globulin (Cytogam, CSL Behring) and a point dilution of palivizumab (Synagis) were included on each plate as positive and negative controls, respectively, and as an assessment of inter-plate variability. All samples were run in duplicate, and results are reported as the average of both replicates. Any sample with an OD value over 1.0 and a CV over 20%, samples with OD between 0.2 and 1.0 with a CV over 30%, or any serial dilution with more than 4 CVs over 20% were repeated one time. Samples that did not meet QC criteria on either biological replicate were excluded. Multiple testing correction was applied to gB AD-2 and gB AD-6 ELISA, but was not applied to post hoc prefusion-like gB, UL16, and gB AD-4+5 assays that were performed to confirm associations identified in our multiplex binding assay.

#### *Binding to HCMV Virions*

Binding to HCMV virions was performed using ELISA in the 384 well format as described above and reported previously[7]. Plates were coated with 100 PFU/well of the AD169r strain at 4°C overnight, blocked, and incubated with three-fold, eight-point serial dilutions for each sample at a 1:20 starting dilution in assay diluent. To calculate the avidity index, primary sample incubation was followed by a seven minute incubation with either PBS or 7M urea. $ED_{50}$ was calculated using the drm and ED functions from the drc package in R. Avidity Index was calculated as the ratio of Urea $ED_{50}$/PBS $ED_{50}$ multiplied by 100.

A serial dilution of HCMV hyperimmuneglobulin (Cytogam, CSL Behring) and a point dilution of palivizumab (Synagis) were included on each plate as positive and negative controls, respectively, and as an assessment of inter-plate variability. All samples were run in duplicate, and results are reported as the average of both replicates. Samples with an OD value over 1.0 and a CV over 20%, samples with OD between 0.2 and 1.0 with a CV over 30%, or any serial dilution with more than 4 CV's over 20% were repeated one time. Samples that did not meet QC criteria on either biological replicate were excluded.

#### *Neutralization Assays*

To determine the capacity of each plasma sample to reduce viral infection, a neutralization assay was performed as described previously[7,8]. Retinal pigment epithelia cells (ARPE-19, ATCC) and Human foreskin fibroblasts (HFF-1, ATCC) were seeded 6,000 cells per well in a 384-well dish (3764, Corning) and incubated overnight at 37°C. Plasma samples were diluted 1:20 and then serially diluted 3-fold with media. Viruses were added at an optimized multiplicity of infection (MOI) (AD169r MOI = 1; Towne MOI = 3). Rabbit complement (Cedarlane) was added to the Towne strain media at an eight-fold dilution. Samples were incubated at 37°C with virus for 1 hour before being transferred to ~90% confluent cells and returned to a 37°C. After 24 hours for HFF-1 cells and 48 hours for ARPE-19 cells, plates are fixed with 10% formalin, washed with a buffer made of DPBS, 10% FBS, and 3% Triton-X-100, and stained with IE1 primary antibody, Goat Anti-Mouse IgG H&L (Alexa Fluor® 488), and 4',6-diamidino-2-phenylindole (DAPI) to detect cells. Plates were imaged and read on the ImageXpress Pico Automated Cell Imaging System (Molecular Devices), which recorded total number of cells and percent infected cells (AF488-positive). The 50% inhibitory dose ($ID_{50}$) was calculated using the drm and ED functions from the drc package in R.

A serial dilution of HCMV hyperimmuneglobulin (Cytogam, CSL Behring) and a point dilution of palivizumab (Synagis) were included on each plate as positive and negative controls, respectively, and as an assessment of inter-plate variability. All samples were run in duplicate, and results are reported as the average of both replicates. Any sample dilution with a % infectivity value over 5.0% and a CV over 20%, samples with a % infectivity value between 1% and 5% with a CV over 30%, or any serial dilution with more than 4 CVs over 20% were repeated one time. Samples that did not meet QC criteria on either biological replicate were excluded.

#### *Inhibition of cell-associated viral spread*

The ability of plasma antibodies to prevent viral cell-associated spread was assessed as described previously[7]. ARPE-19 cells were seeded 5,000 cells per well in a 384 well dish (3764 Corning) and infected approximately 24 hours later with Ts15nr (epithelial-tropic Towne-BAC-GFP with intact pentamer, MOI = 0.05), except for cell only control wells. After 48 hours, cells were washed with PBS fortified with 1% FBS. Samples were diluted in a 96 well round bottom dish 1:20 with ARPE-19 media (DMEM-F12, 10% FBS, 50 U/ml penicillin and streptomycin). Diluted samples were added to the cells in duplicate. Cells were incubated with serum for 12 days at 37°C. After 12 days cells were fixed and analyzed on the ImageXpress Pico Automated Cell Imaging System (Molecular Devices) as described for neutralization assays above. The % spread relative to control was calculated as (% average infection for samples – % average infection for virus-only control)/ % average infection for virus-only control.

A serial dilution of HCMV hyperimmuneglobulin (Cytogam, CSL Behring) and a point dilution of palivizumab (Synagis) were included on each plate as positive and negative controls, respectively, and as an assessment of inter-plate variability. All samples were run in duplicate, and results are reported as the average of both replicates. Any sample dilution with a % infectivity value over 5.0% and a CV over 20%, samples with a % infectivity value between 1% and 5% with a CV over 30%, or any serial dilution with more than 4 CVs over 20% were repeated one time. Samples that did not meet QC criteria on either biological replicate were excluded.

#### *Antibody Dependent Cellular Phagocytosis*

An optimized titer of AD169r virions (125 PFU/well) or Towne virions (1250 PFU/well) was conjugated to Alexa Fluor 647 (AF647) N-hydroxysuccinimide (NHS) ester prior to incubation with diluted sera samples (1:20). Then, virus-antibody immune complexes were centrifuged with 50,000 THP-1 cells for 1 hour at 1,200 ×g and incubated at 37°C for 1 hour. Cells were stained with Aqua Live/Dead stain, fixed with 10% formalin, and washed prior to acquisition on the flow cytometer (Fortessa; BD) using the HTS as previously described [7,8]. The percentage of AF647+ cells was reported for each sample based on the live, singlet population, and the average %AF647+ cells of the PBS wells was subtracted from the %AF647+ cell values of controls and study samples for background correction. A positivity threshold was calculated as the average + 2 standard deviations of two seronegative plasma samples.

A serial dilution of HCMV hyperimmuneglobulin (Cytogam, CSL Behring) and a point dilution of palivizumab (Synagis) were included on each plate as positive and negative controls, respectively, and as an assessment of inter-plate variability. All samples were run in duplicate, and results are reported as the average of both replicates. Samples with viability <85%, event count below 4000, or a CV greater than 30% were repeated one time. Samples that did not meet QC criteria on either biological replicate were excluded.

#### *Binding to cell-associated glycoprotein B*

Binding to the Merlin strain of gB expressed on the surface of the cell was measured as described previously[7,9]. Briefly, plasmids expressing GFP and gB were co-transfected into HEK293T cells for 48 hours at 37°C. Transfected cells were then incubated with plasma diluted 1:6,250. Cells were stained with Live/Dead Fixable Near-IR Dead Cell Stain, followed by phycoerythrin (PE)-conjugated goat-anti-mouse IgG Fc staining, and fixed with 10% formalin prior to acquisition via high throughput sampler (HTS) on the flow cytometer (Fortessa; BD). The percentage of PE+ cells was reported for each sample based on the live, singlet, GFP+ population. A positivity threshold was calculated as the average + 2*SD of two seronegative plasma samples. A serial dilution of HCMV hyperimmuneglobulin (Cytogam, CSL Behring) and a point dilution of palivizumab (Synagis) were included on each plate as positive and negative controls, respectively, and as an assessment of inter-plate variability. All samples were run in duplicate, and results are reported as the average of both replicates. Samples with viability lower than 90%, event count below 8000, or a CV greater than 30% were repeated one time. Samples that did not meet QC criteria on either biological replicate were excluded.

#### *HCMV viral load in plasma samples*

DNAemia was measured vis HCMV IE1-specific qPCR as previously reported[10]. Viral DNA was extracted using High Pure Viral Nucleic Acid Extraction Kit (Roche) from 200 mcL of plasma with a final elution volume of 50 mcL. For the PCR reaction 5 mcL of sample was added to duplicate wells containing 25 mcL mixture of 15 mcL of SybrSelect (Invitrogen), 300 nM of primers (Integrated DNA Technologies) and water. Primers were designed to the immediate early 1 (IE1) gene as previously described.1 qPCR conditions consisted of a two minute cycle at 50°C, followed by 10 minutes at 95°C, 40 cycles of denaturation at 95°C for 15 seconds and combined annealing/extension at 63°C for 1 minute. Serial dilutions of pre-quantitated AD169 virus (Advanced Biotechnologies) with a range of 1-$10^5$ copies per mL were run in triplicate to create a standard curve. Virus was considered detected at >100 copies per mL. The sample was considered positive for HCMV by qPCR if at least 2 out of 6 replicates had virus detected. Samples were repeated until 2 replicates were positive or 6 replicates were completed, whichever came first. Replicates that were undetected or below 100 copies/mL were set to 50 copies/mL. Viral loads for positive samples were calculated by averaging all replicates, including negative replicates.

#### *Packages used for statistical analysis*

Case-control matching based on the criteria described in the main text was performed using the Matching package (version 4.10-14) in R[11]. To assess whether an immune marker was associated with our transmission, we performed conditional logistic regression using the survival package[12]. Heatmaps were made using the complexHeatmap package in R[13]. Test of the associated of DNAemia with immune marker was performed using the coin package[14]. Conditional logistic LASSO was performed using the clogitL1 package[15]. Multiple imputation to address missing data in the LASSO analysis was performed using the mice package with the maximum iterations set to 5[16].

#### *Sensitivity Analyses*

While all care was taken to run matches on the same plate and date while measuring immune functions, repeated samples due to QC failure may not have been on the same plate as their matched participants. To address this, we repeated the conditional logistic regression restricted to matches run on the same plate to ensure results were not driven by between inter-plate variability. In addition, the conditional logistic regression was rerun for significant immune markers adjusting for presence of DNAemia (when available) to see if presence of DNAemia impacted our results (Table S3). The significance for all markers did not change after either of these sensitivity analyses. Sensitivity analyses of the conditional logistic LASSO are described in the main text (Figures S12-S14).

#### *Statement on the use of generative AI*

During the preparation of this work, the authors used Microsoft Copilot and Google Gemini to optimize code for figure and table generation. The authors reviewed and edited the suggested code and take full responsibility for all content. No generative AI was used for the drafting or editing of this manuscript.

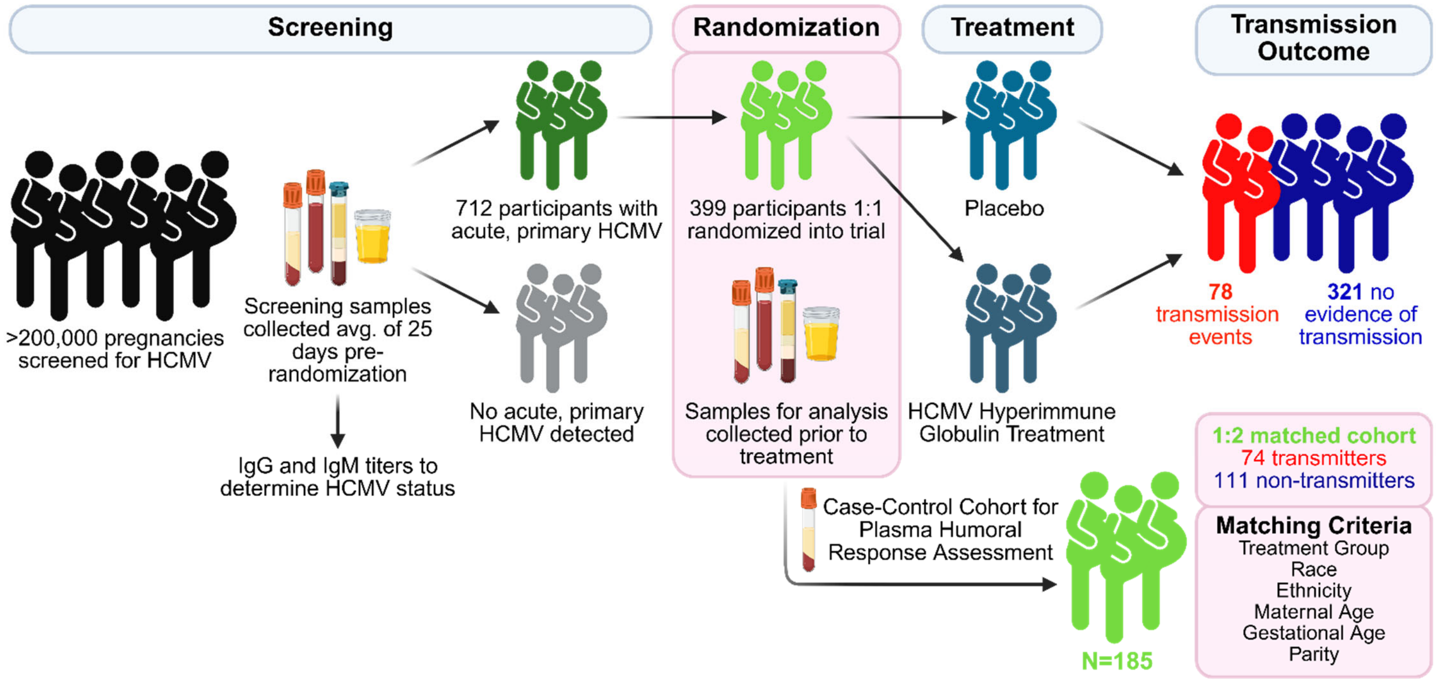

**FIGURE S1. STUDY OVERVIEW**

Over 200,000 pregnancies were screened as part of A Randomized Trial to Prevent Congenital Cytomegalovirus (CMV)(NCT01376778)[1]. Pregnant women were eligible for serological screening for primary HCMV infection if they had a singleton pregnancy prior to 23 weeks gestation. Primary infection was defined as positive IgM and low avidity IgG or IgG seroconversion. Samples for this study were collected at the randomization timepoint, prior to administration of the hyperimmune globulin or placebo, which occurred 24.56± 8.45 (mean ± SD) days post screening. Of the 399 women who underwent randomization, 78 participants had documented transmission events. Transmission was defined as fetal loss with pathologic evidence of HCMV infection or detection in amniotic fluid and/or infant urine or saliva prior to 21 days of life. For the immune correlate study, plasma was processed from collected whole blood. For immune marker analysis, cases (women with a confirmed transmission event) and controls (women without confirmed transmission) were matched on demographic features that could impact HCMV exposure: treatment group (HIG treatment or placebo), race (White, Black, Other/Mixed/Not reported), ethnicity (Hispanic/Latino or Non-Hispanic/Latino), maternal age (within 10 years), gestational age (within 5 weeks), and parity (nulliparous or multiparous)(Table 1). 71 cases were matched with two controls, while three cases were matched with one control. Due to sample size, 22 controls were matched to more than one transmitter. Four cases did not have any suitably matched controls and were excluded from analysis.

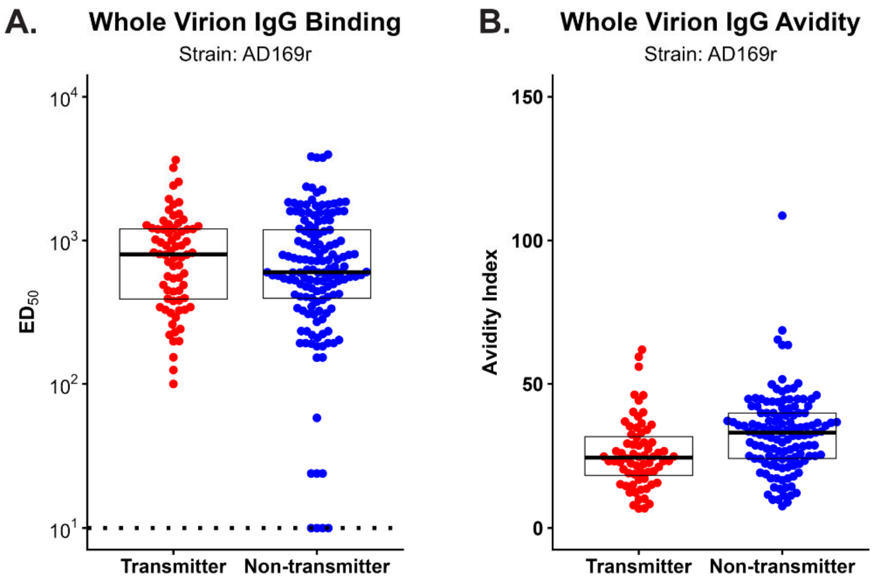

**FIGURE S2. BINDING TO HCMV STRAIN AD169r**

IgG binding (A) and avidity (B) to HCMV virions measured via ELISA in transmitters (red) and non-transmitters (blue). Participants without detectable binding were set to $ED_{50}$=10 (dashed line), which is half of the starting dilution (1:20). $ED_{50}$ was calculated using the drm package in R. Avidity index calculated as the ratio of binding in the presence of urea to binding in the presence of PBS. Conditional logistic regression was used to assess association with transmission odds for binding (p=0.2, q=0.2) and avidity (p<0.001, q<0.001).

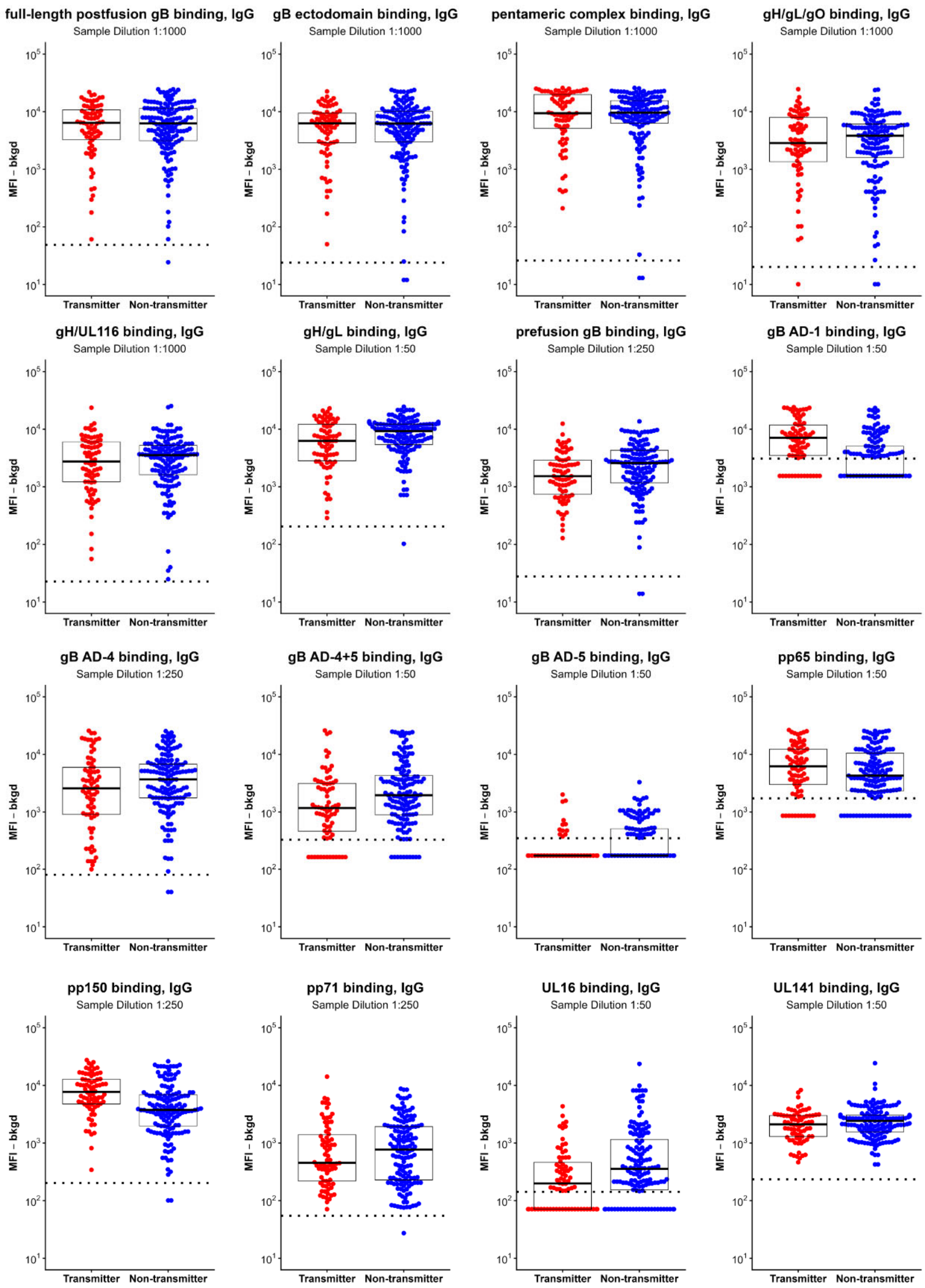

**FIGURE S3. ANTIGEN-SPECIFIC PLASMA IgG BINDING**

IgG binding to 16 HCMV antigens was measured via multiplex-binding assay in transmitters (red) and non-transmitters (blue) and reported as the background subtracted median fluorescence intensity (MFI-bkgd). Positivity threshold (dotted line) for each antigen was calculated as the average + 2*SD of a panel of seronegative plasma samples. Participants with MFI-bkgd less than the positivity threshold were set to half of the positivity threshold. Conditional logistic regression was used to assess association with transmission odds for each antigen (Table S2).

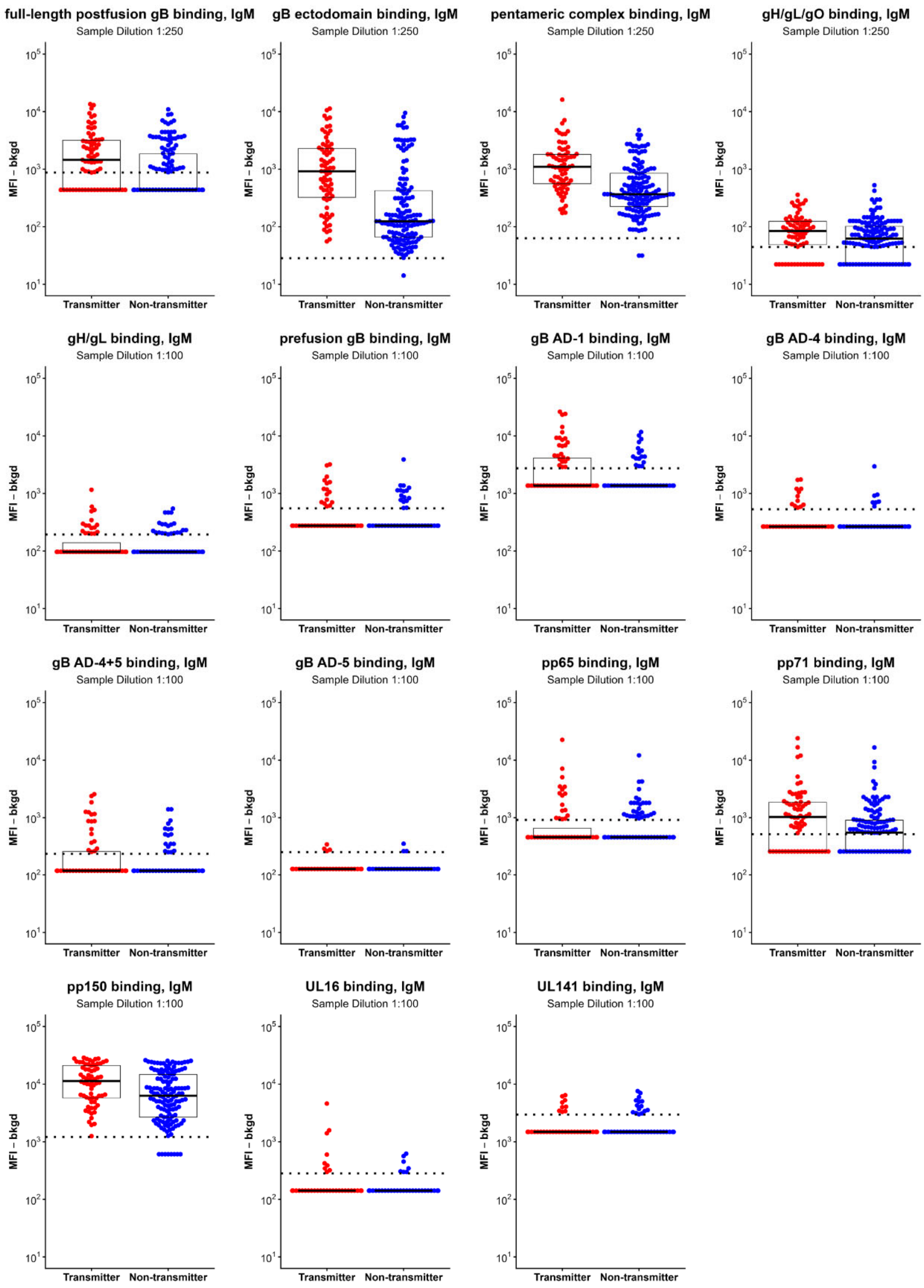

**FIGURE S4. ANTIGEN-SPECIFIC PLASMA IgM BINDING**

IgM binding to 15 HCMV antigens was measured via multiplex-binding assay in transmitters (red) and non-transmitters (blue) and reported as the background subtracted median fluorescence intensity (MFI-bkgd). Positivity threshold (dotted line) for each antigen was calculated as the average + 2*SD of a panel of seronegative plasma samples. Participants with MFI-bkgd less than the positivity threshold were set to half of the positivity threshold. Conditional logistic regression was used to assess association with transmission odds for each antigen (Table S2).

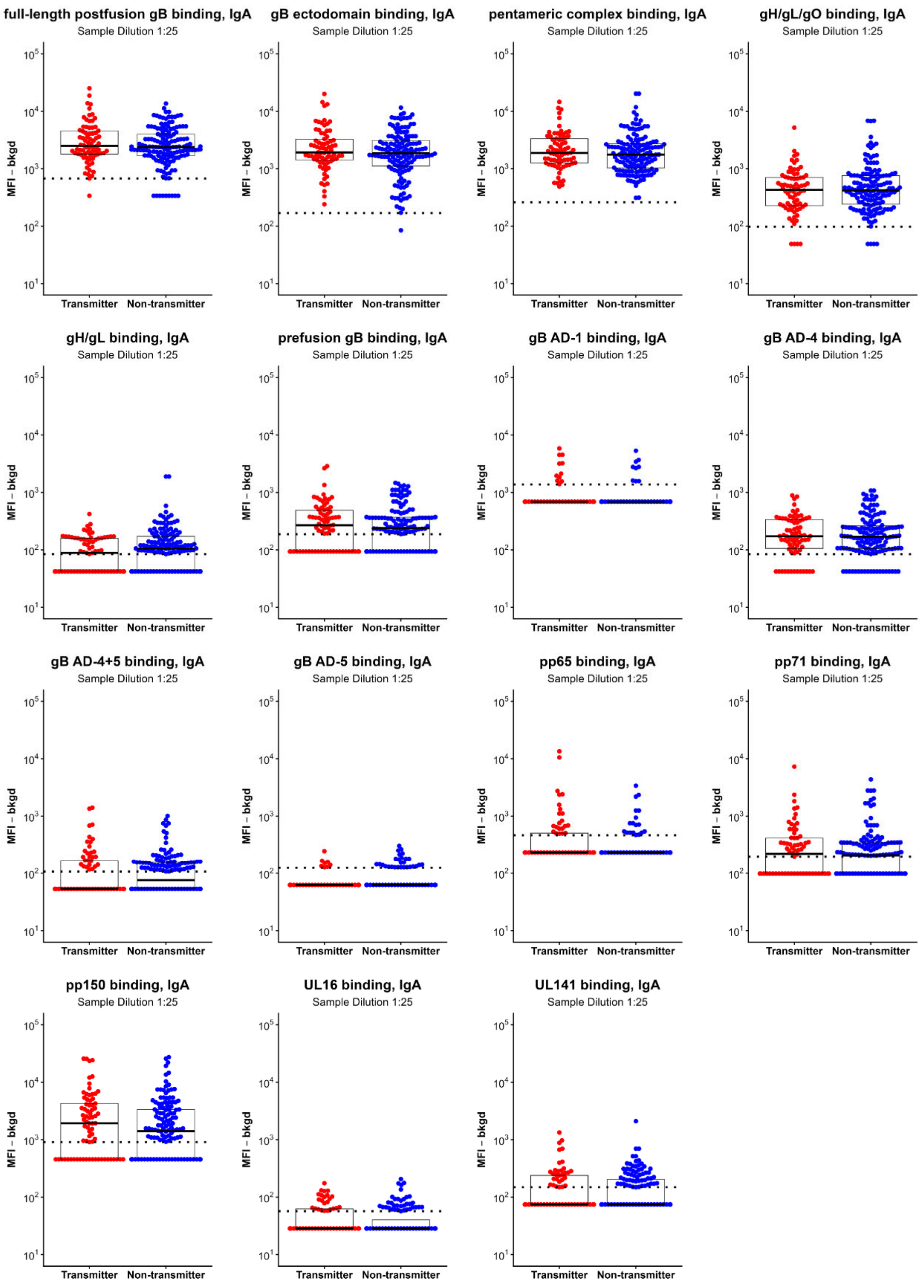

**FIGURE S5. ANTIGEN-SPECIFIC PLASMA IgA BINDING**

IgA binding to 15 HCMV antigens was measured via multiplex-binding assay in transmitters (red) and non-transmitters (blue) and reported as the background subtracted median fluorescence intensity (MFI-bkgd). Positivity threshold (dotted line) for each antigen was calculated as the average + 2*SD of a panel of seronegative plasma samples. Participants with MFI-bkgd less than the positivity threshold were set to half of the positivity threshold. Conditional logistic regression was used to assess association with transmission odds for each antigen (Table S2).

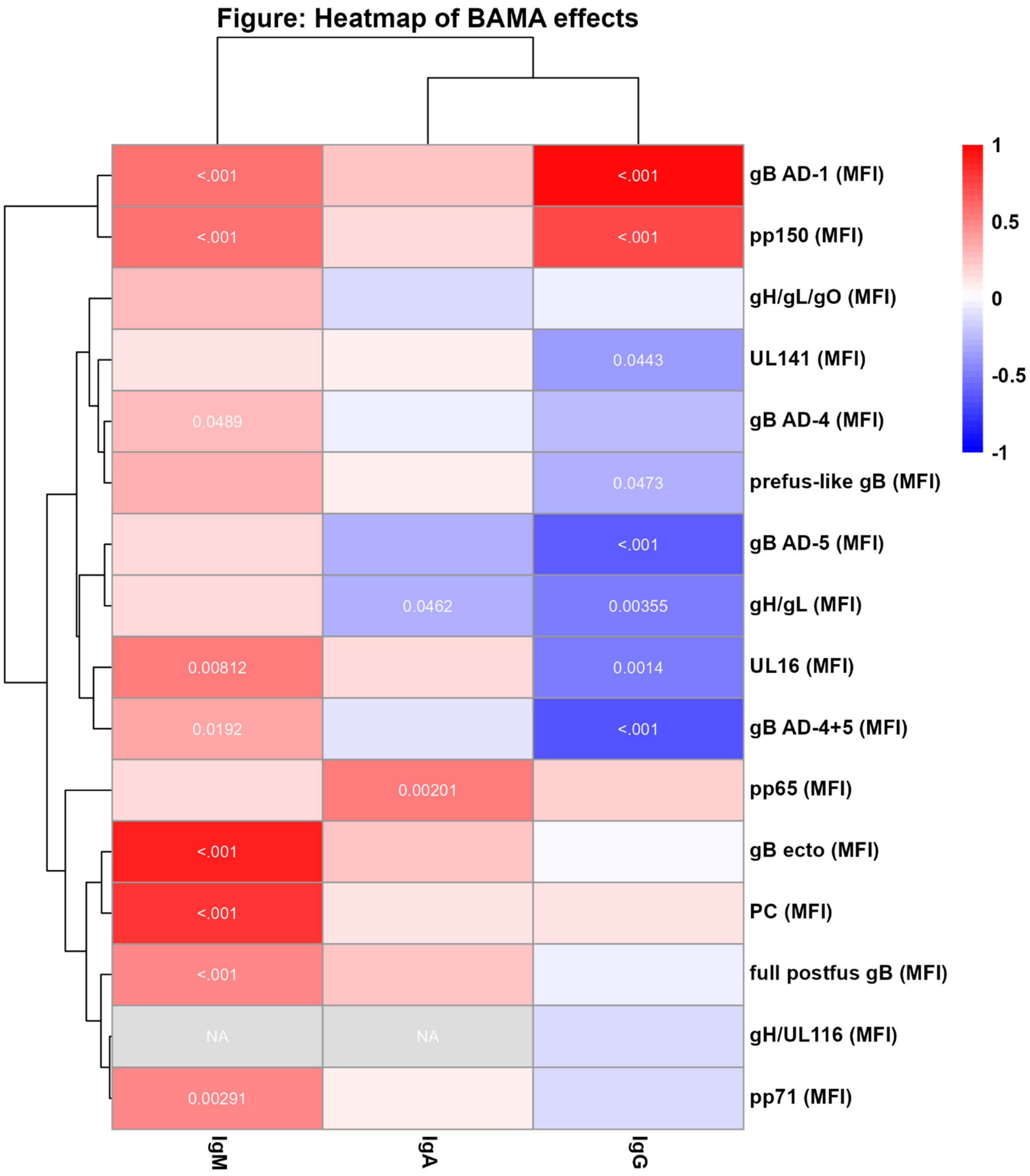

**FIGURE S6. HEATMAP OF ANTIGEN-SPECIFIC PLASMA ANTIBODY BINDING**

Heatmap of the association of IgG (Figure S3), IgM (Figure S4), and IgA (Figure S5) binding with transmission odds. Coloring scale indicates the log odds ratio for each response with red indicating increased odds of transmission and blue indicated decreased odds of transmission. Unadjusted p-value displayed for markers with conditional logistic regression p-value <0.05. Hierarchical clustering performed using heatmap package in R.

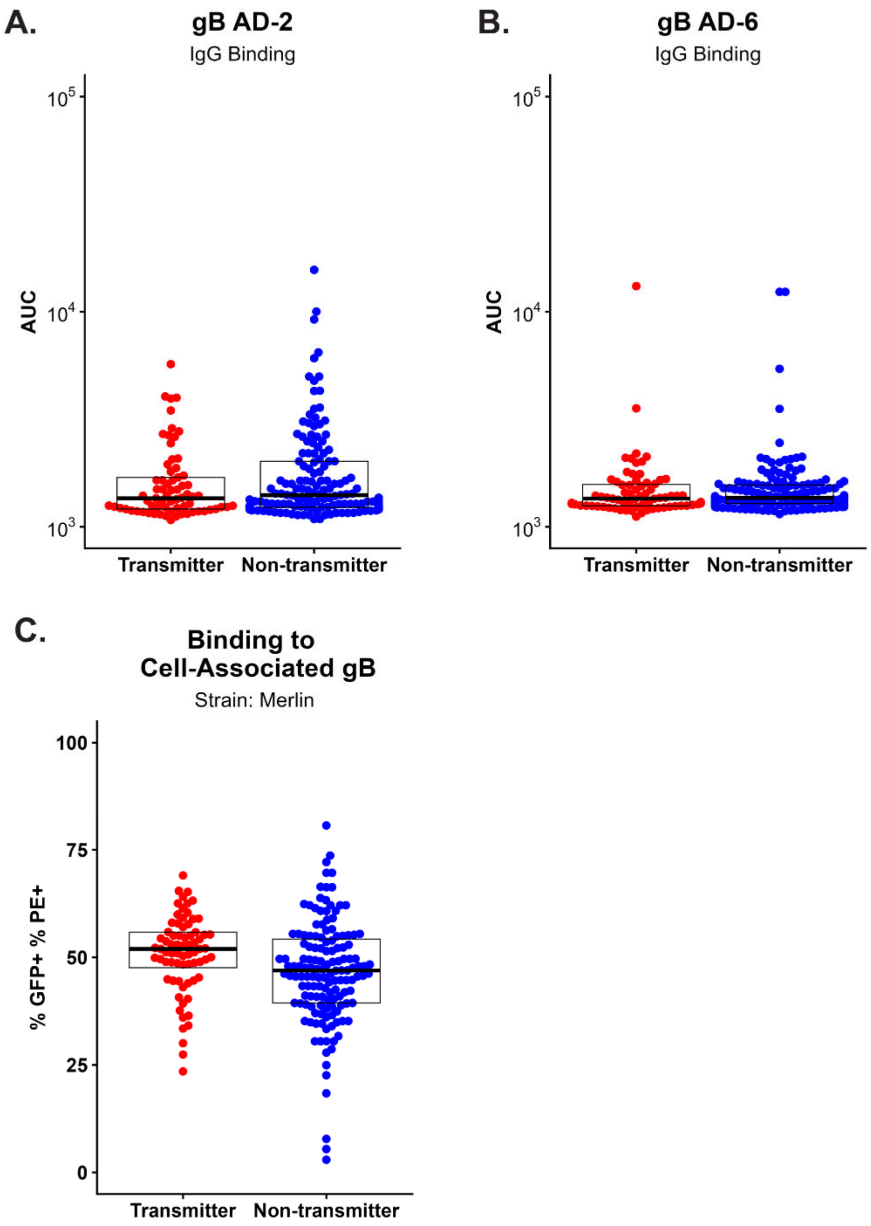

**FIGURE S7. IgG BINDING TO gB AD-2, gB AD-6, AND CELL-ASSOCIATED gB**

(A) IgG binding to gB AD-2 (p=0.16, q=0.31) and gB AD-6 (p=0.75, q=0.75) in transmitters (red) and non-transmitters (blue) was measured via ELISA and reported as area under the curve. Plasma samples were diluted 1:10 and serially diluted three-fold. Area under the curve for each participant was calculated using the AUC function from the DescTools package in R. (B) IgG binding to cell-associated gB (p=0.005) in transmitters (red) and non-transmitters (blue) was measured via transfected cell assay. Cells were co-infected with GFP and gB expressing plasmids. IgG is detected with phycoerythrin-conjugated secondary antibody. Cells were analyzed via flow cytometry and reported as the percentage of cells positive for GFP and PE.

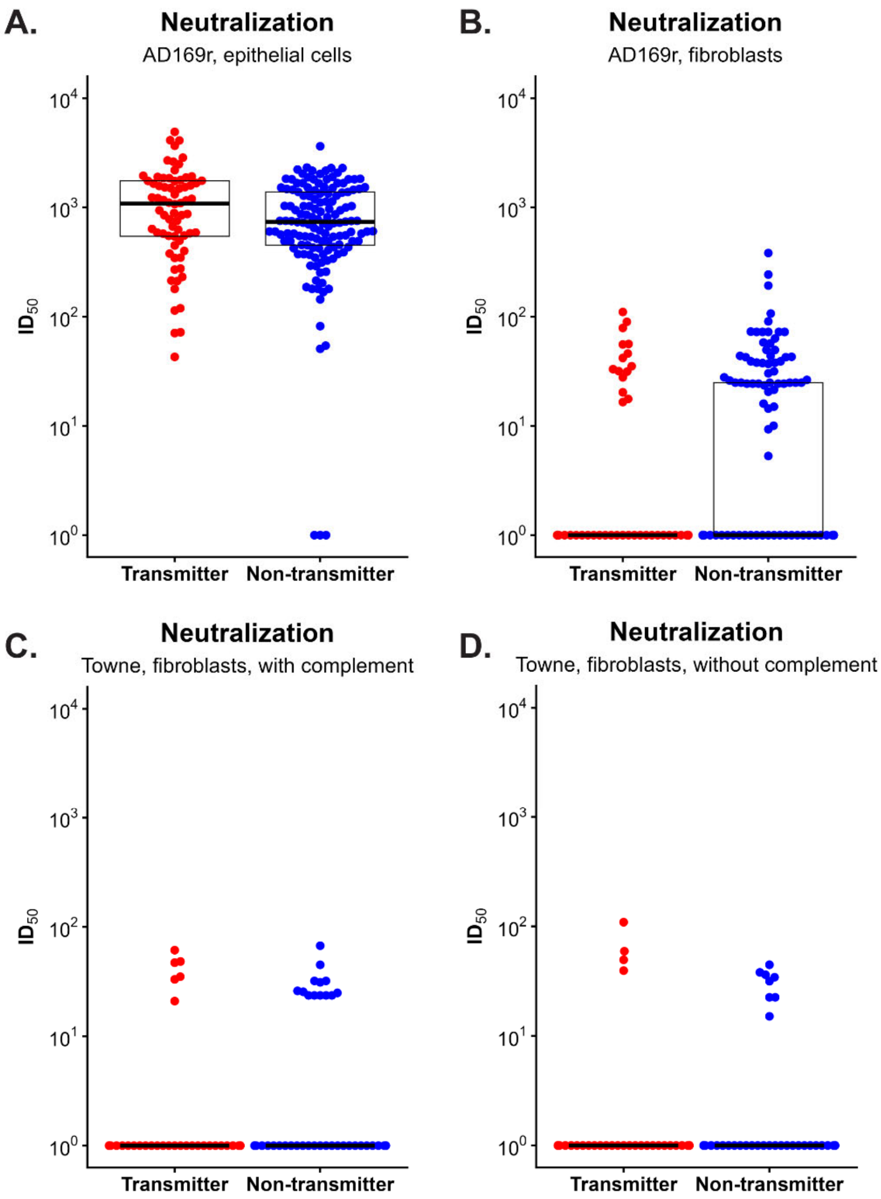

**FIGURE S8. NEUTRALIZATION OF HCMV**

HCMV neutralization was measured for transmitters (red) and non-transmitters (blue) using the AD169r strain on epithelial (A)($p=0.058$, $q=0.087$) and fibroblast (B)($p=0.017$, $q=0.052$) cells, and with the Towne strain on fibroblasts with (C) and without (D) complement. Neutralization is reported as $ID_{50}$. Participants with less than 50% reduction in viral infection for the starting dilution were set to $ID_{50}=0$ for analysis and $ID_{50}=1$ for visualization on a logarithmic scale.

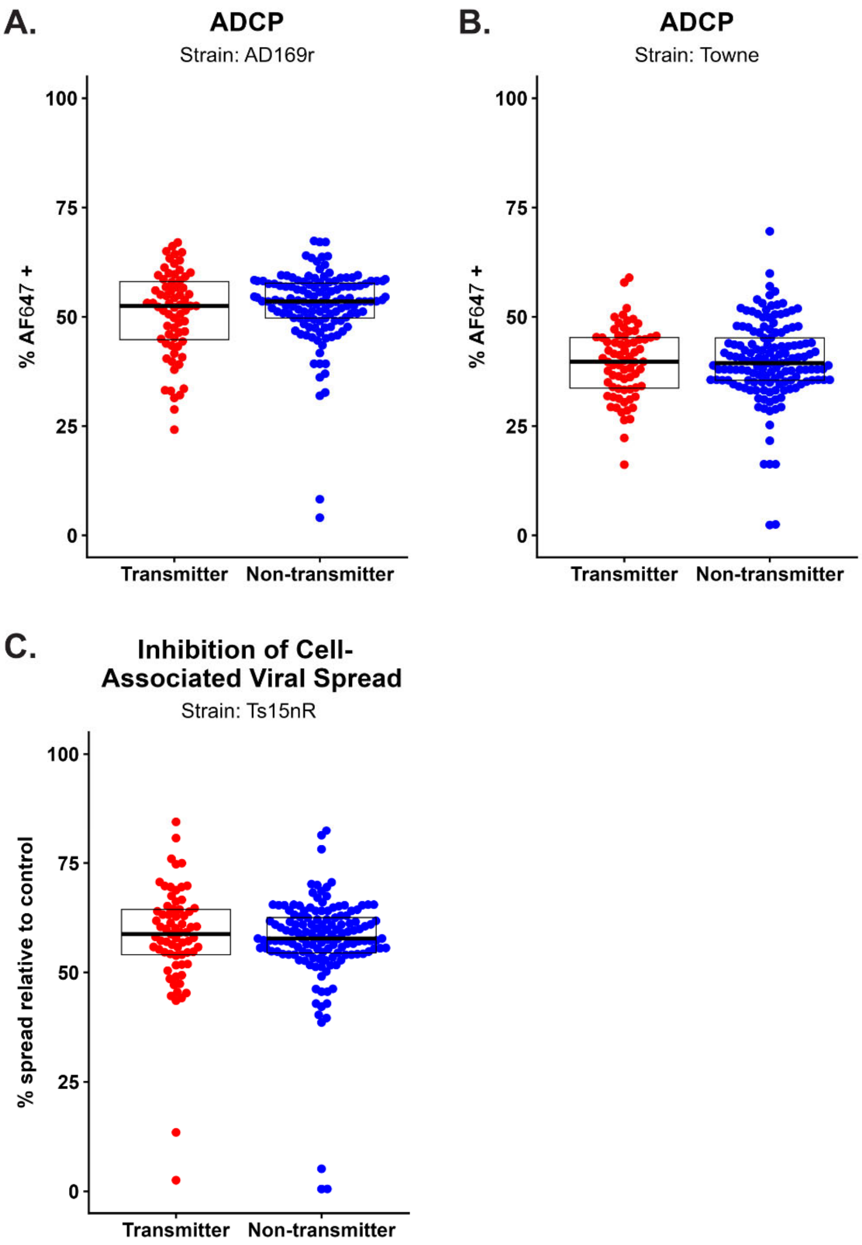

**FIGURE S9. NON-NEUTRALIZING FUNCTIONAL RESPONSES**

Antibody-dependent cellular phagocytosis was measured for transmitters (red) and non-transmitters (blue) using AF647+-conjugated AD169r (A)(p=0.14, q=0.28) or Towne (B)(p=0.75, q=0.75) virus on the THP-1 monocyte cell line and reported as %AF647+ cells. Inhibition of cell-associated viral spread (C)(p=0.96) was measured using the reduction of cell-associated spread of the Ts15nR strain in the presence of plasma diluted 1:10.

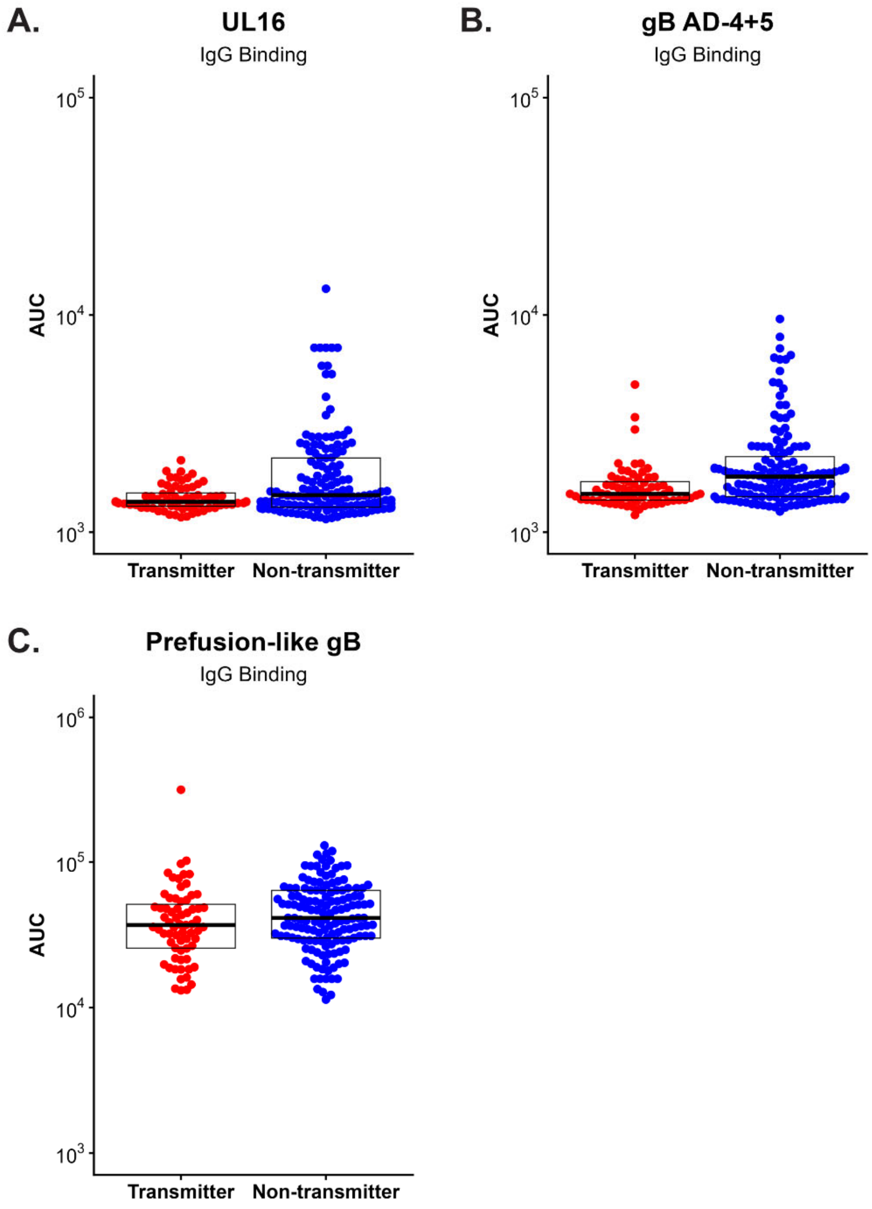

**FIGURE S10. POST HOC IgG BINDING ELISA**

We measured binding via ELISA to three antigens that were significantly associated with reduced odds in transmitters (red) and non-transmitters (blue) in our semi-quantitative multiplex binding assay. IgG binding to UL16 (A)($p<0.001$), gB AD-4+5 (B)($p<0.001$), and prefusion-like gB (C)($p=0.11$) is reported as area under the curve. Multiple testing correction was not performed for post hoc assays. Area under the curve for each participant was calculated using the AUC function from the DescTools package in R.

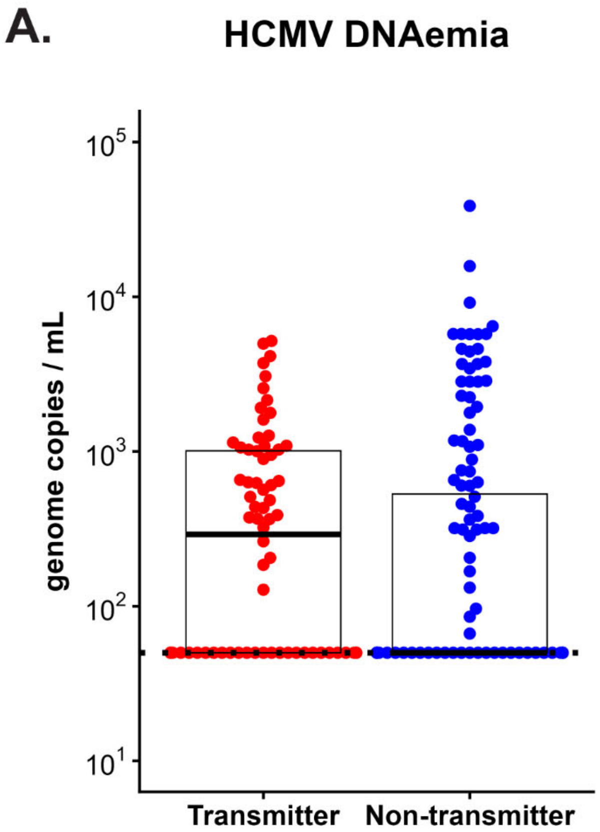

**FIGURE S11. HCMV DNAEMIA AT THE TIME OF SAMPLE COLLECTION**
DNAemia was measured via IE1-specific qPCR and reported as genome copies/mL. Participants were considered positive if they had detectable DNA >100 copies/mL in at least 2 of 6 replicates. DNAemia is reported as the average of all positive replicates. Participants without detectable DNAemia were set to 50 copies/mL, which is half of the positivity threshold. Conditional logistic regression to assess association with transmission odds between participants with and without detectable DNAemia (p=0.005).

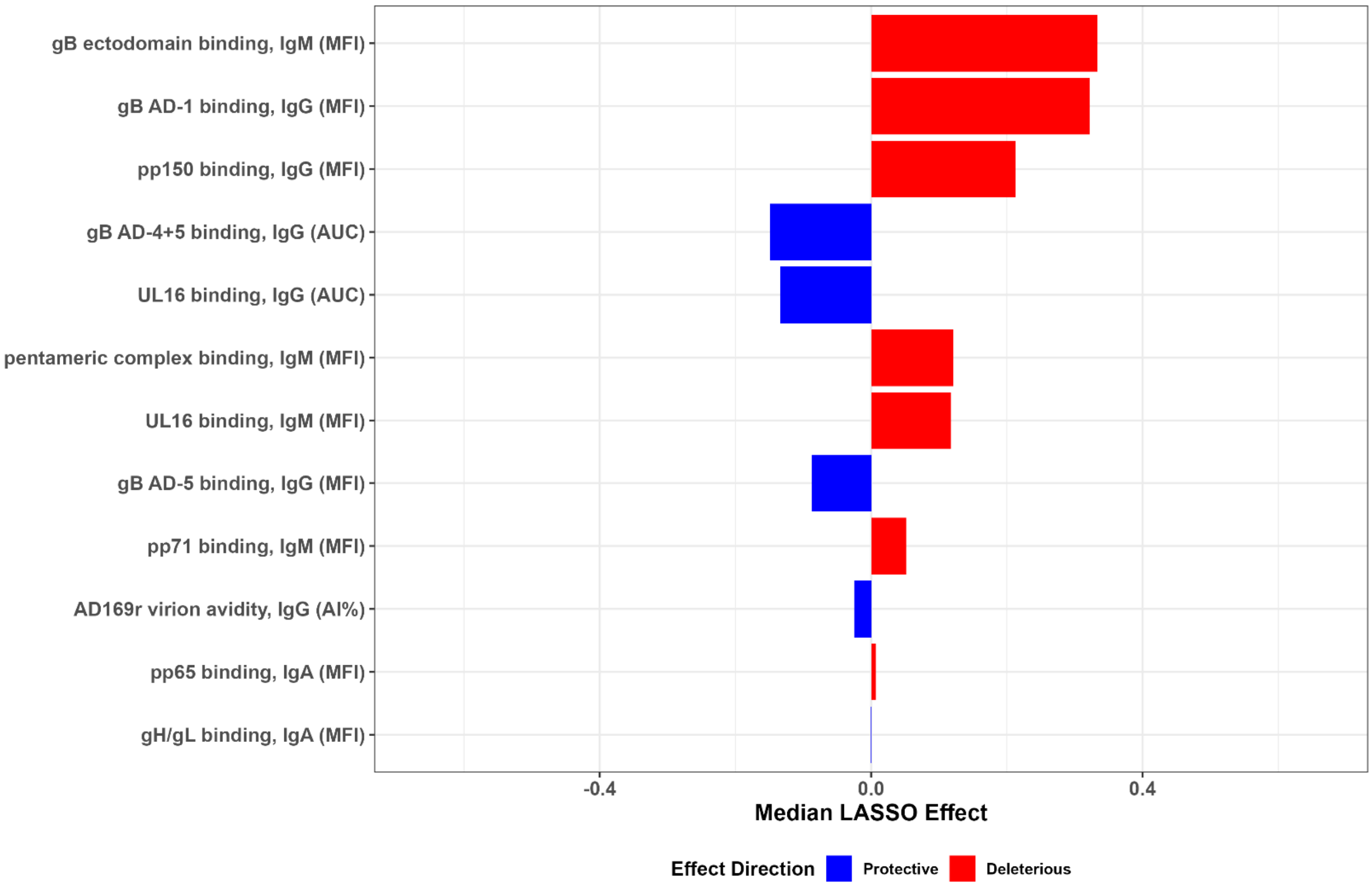

**FIGURE S12. CONDITIONAL LOGISTIC LASSO WITH CORRELATION PRUNING**
Conditional logistic LASSO (Least Absolute Shrinkage and Selection Operator) was performed using all participants with multiple imputations for missing data. Here, the analysis was repeated with variables with Spearman r>0.8 removed to eliminate bias from highly correlated variables.

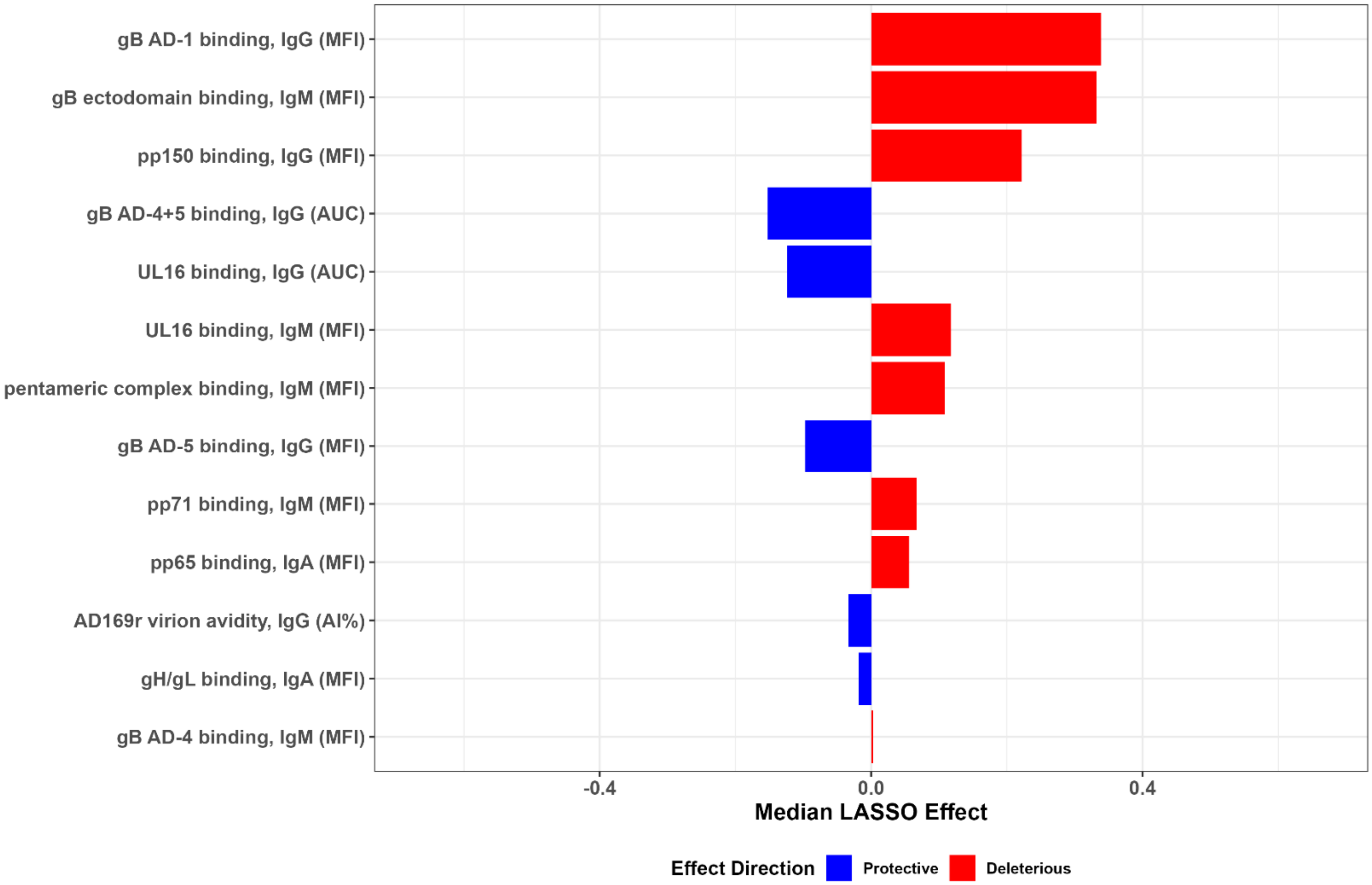

**FIGURE S13. CONDITIONAL LOGISTIC LASSO INCLUDING DNAEMIA AS A VARIABLE**
Conditional logistic LASSO (Least Absolute Shrinkage and Selection Operator) was performed using all participants with multiple imputations for missing data. Here, the analysis was repeated including presence of DNAemia as a variable. Presence of DNAemia was not selected in the model.

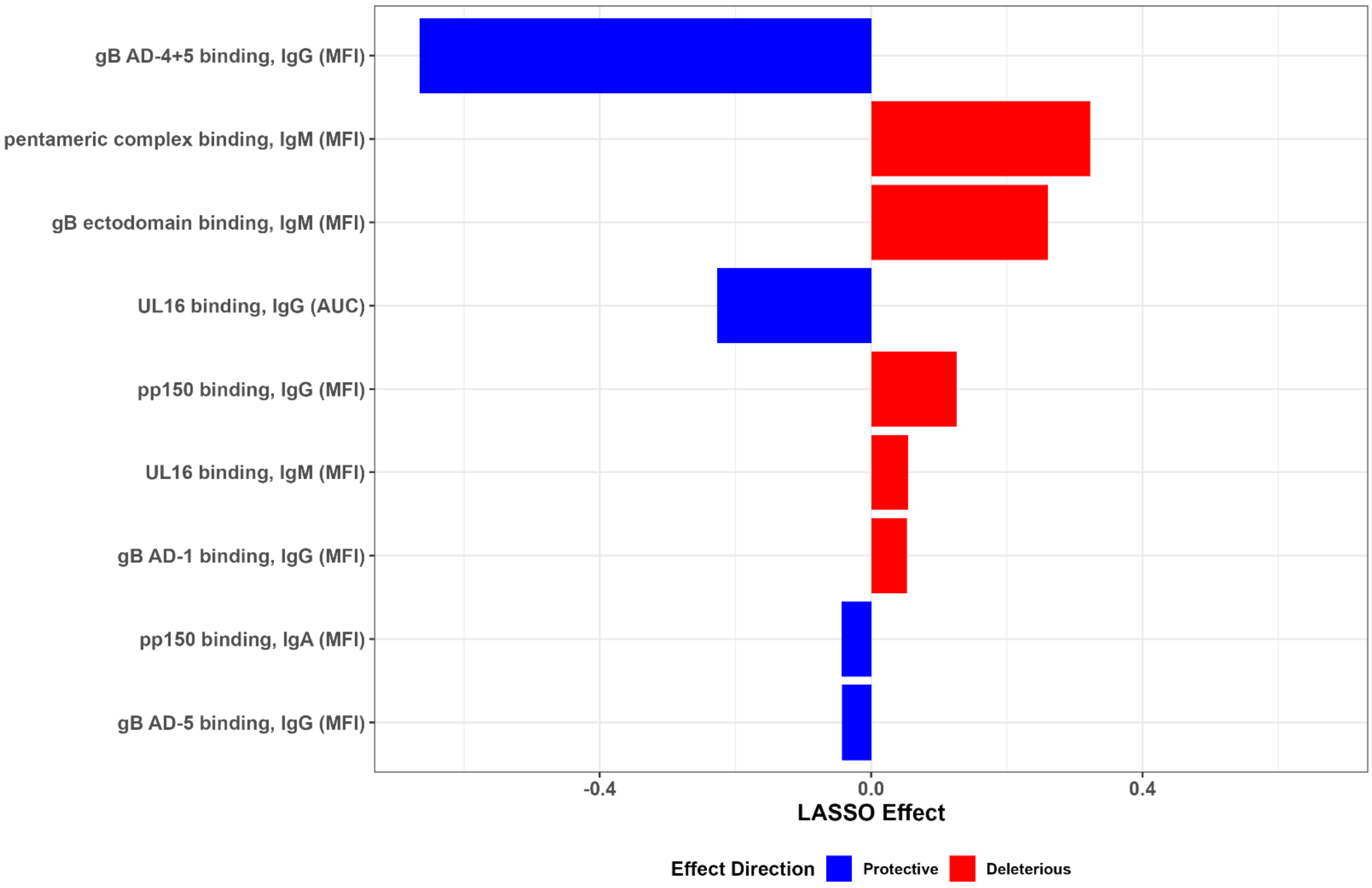

**FIGURE S14. CONDITIONAL LOGISTIC LASSO RESTRICTED TO PARTICIPANTS WITH COMPLETE DATA**

Conditional logistic LASSO (Least Absolute Shrinkage and Selection Operator) was performed using all participants. Here, the analysis was repeated without multiple imputation using data from only the n=95 participants with data that passed quality control for all 55 immune markers.

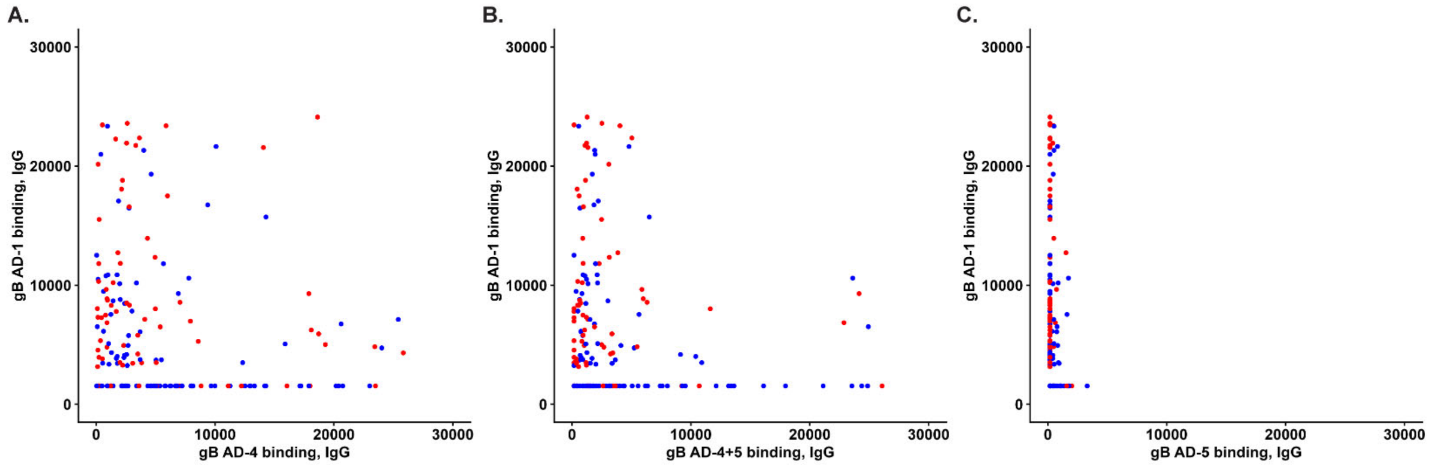

**FIGURE S15. CORRELATION BETWEEN GB ANTIGENIC DOMAIN PLASMA IGG BINDING**

IgG binding to was measured via multiplex-binding assay in transmitters (red) and non-transmitters (blue) and reported as the background subtracted median fluorescence intensity (MFI-bkgd). Correlations are shown between gB AD-1 and gB AD-4 (Spearman ρ = -0.2424), gB AD-4+5 (Spearman ρ= -0.3029), and gB AD-5 (Spearman ρ = -0.1041).

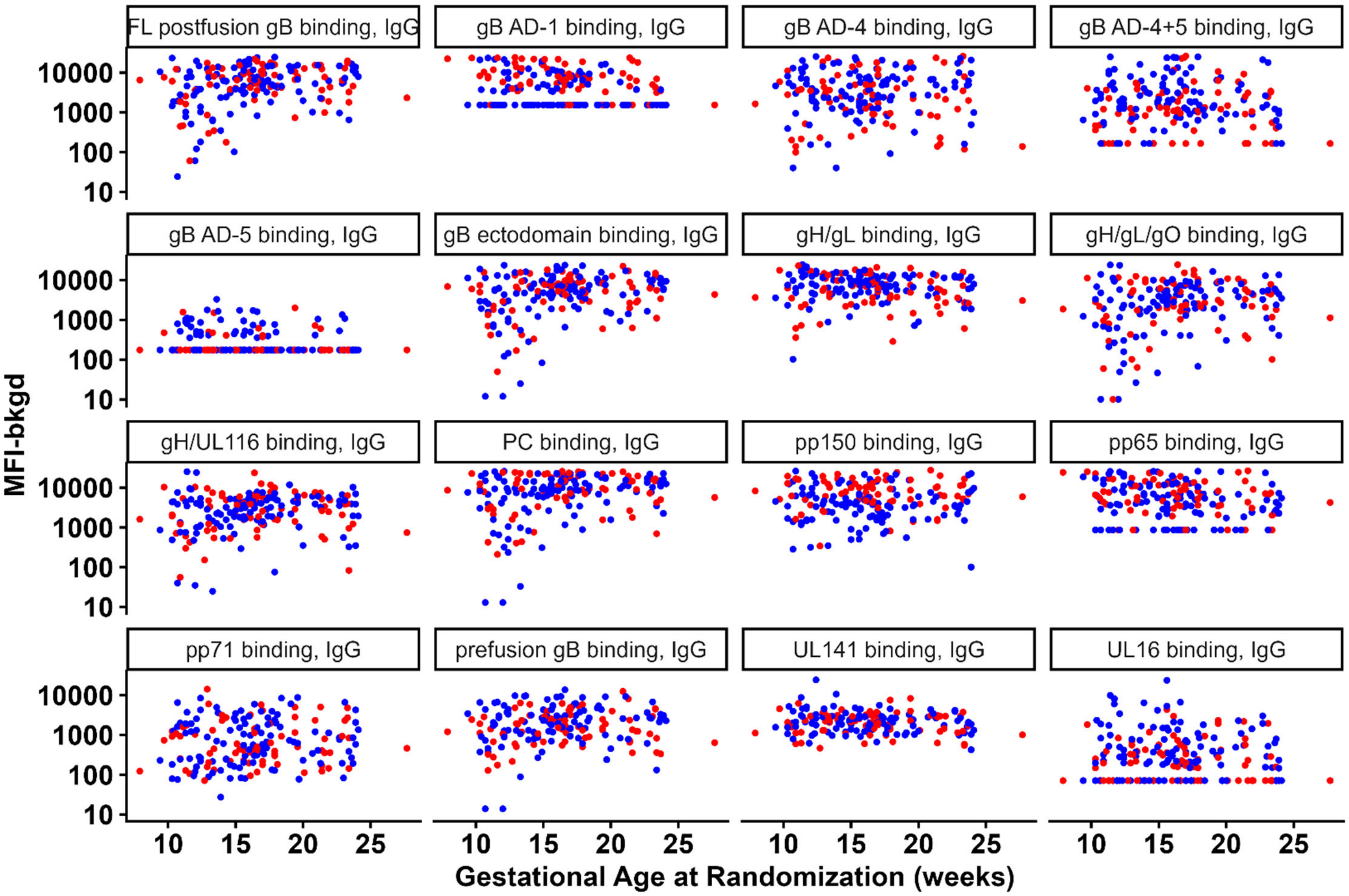

**FIGURE S16. IGG BINDING BY GESTATIONAL AGE AT RANDOMIZATION**

IgG binding to 16 HCMV antigens was measured via multiplex-binding assay in transmitters (red) and non-transmitters (blue) and reported as the background subtracted median fluorescence intensity (MFI-bkgd). Data is graphed by the participant's gestational age at randomization.

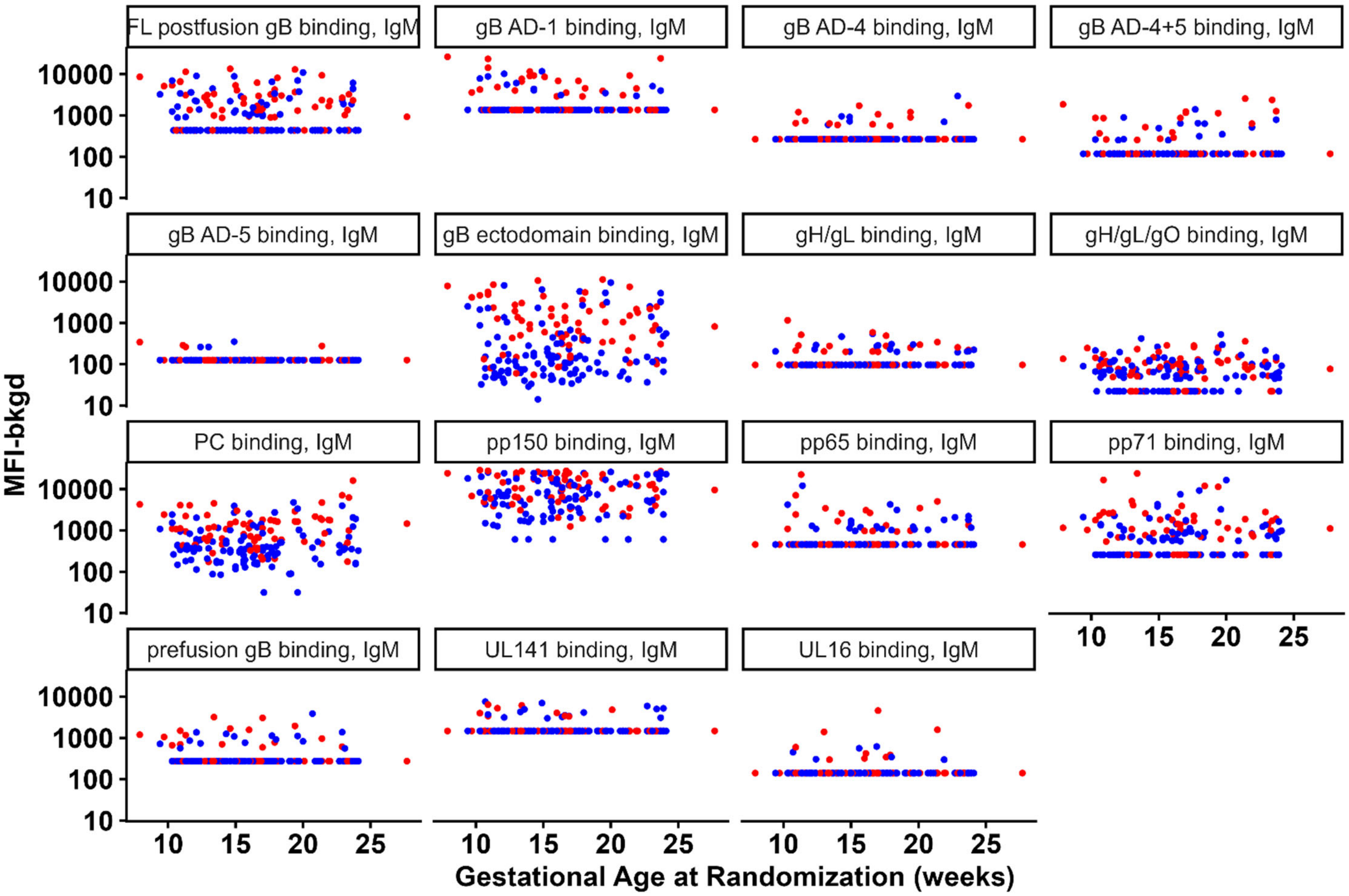

**FIGURE S17. IGM BINDING BY GESTATIONAL AGE AT RANDOMIZATION**

IgM binding to 15 HCMV antigens was measured via multiplex-binding assay in transmitters (red) and non-transmitters (blue) and reported as the background subtracted median fluorescence intensity (MFI-bkgd). Data is graphed by the participant's gestational age at randomization.

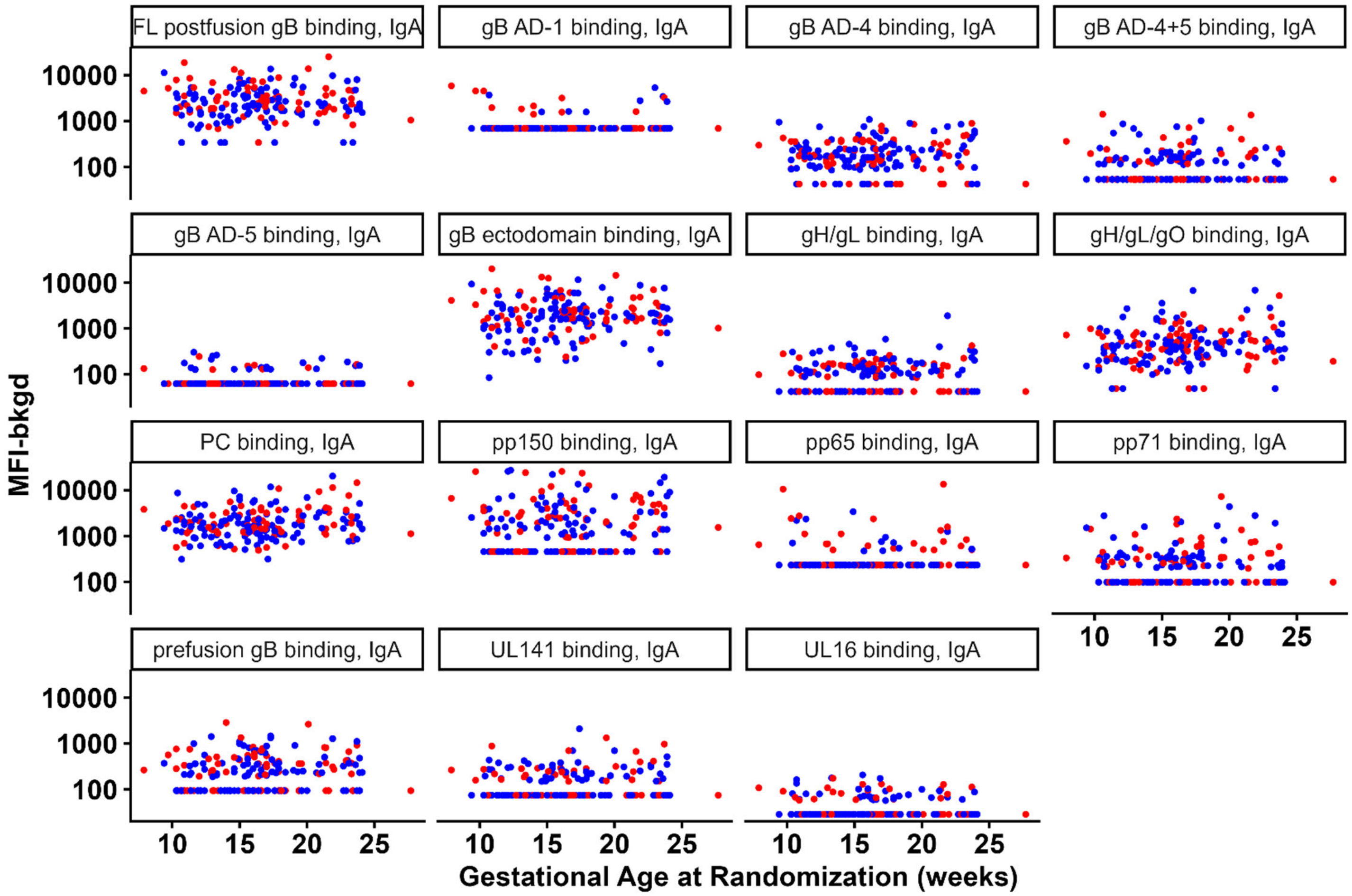

**FIGURE S18. IGA BINDING BY GESTATIONAL AGE AT RANDOMIZATION**

IgA binding to 15 HCMV antigens was measured via multiplex-binding assay in transmitters (red) and non-transmitters (blue) and reported as the background subtracted median fluorescence intensity (MFI-bkgd). Data is graphed by the participant's gestational age at randomization.

**TABLE S1. HCMV ANTIGEN DETAILS**

| Name | Source | Sequence Information |
|---|---|---|
| **full length postfusion gB** | Sino Biological #10202-VCCH1 | Transmembrane domain-deleted sequence of the Towne strain of HCMV gB extracellular domain (AAA45920.1, aa 1-700, 777-907, fused with five additional amino acids (DDDDK) at the C-terminus). Furin cleavage site is mutated from 'RTKR' to 'TTQT'. |
| **gB ectodomain** | Produced in house | Towne strain |
| **Prefusion-like gB** | Kind gift from Dr. Jason McClellan | PDB ID: 8VY004E |
| **gB AD-1** | MyBioSource #MBS485117 | Residues 552–647 of HCMV gB Merlin Strain (UniProtKB: F5HB53) |
| **gB AD-2** | GenScript Custom Peptide | AD169 Strain; HRANETIYNTTLKYG |
| **gB AD-4** | Produced in house | Residues 112–132 and 343–438 of HCMV gB Merlin strain (UniProtKB: F5HB53), Ile-Ala-Gly-Ser-Gly flexible linker |
| **gB AD-4+5** | Produced in house | Residues 112–438 of HCMV gB Merlin strain (UniProtKB: F5HB53 |
| **gB AD-5** | Produced in house | Residues 133–343 of HCMV gB Merlin strain (UniProtKB: F5HB53) |
| **gB AD-6** | GenScript Custom Peptide | Towne strain, CMIALDIDPLENTDFRVLELYSQKELRSSNVFDLEEIMREFNSYKQRVKYV |
| **pentameric complex** | The Native Antigen Company #CMV-PENT | VR1814 strain; GenBank: ACZ79986 |
| **gH/gL/gO** | Produced in house | TB40/E strain |
| **gH/gL** | Produced in house | TB40/E strain |
| **pp65** | abcam #ab43041 | AD169 strain; UniProt: P06725 |
| **pp71** | Produced in house from plasmid; kind gift from Dr. Robert Kaletja | AD169 strain |
| **pp150** | abcam #ab43039 | Merlin strain; UniProt: Q6SW05 |
| **UL16** | Kind gift from Dr. Richard Stanton | Merlin strain |
| **UL141** | Kind gift from Dr. Richard Stanton | Merlin strain |
| **gH/UL116** | Kind gift from Dr. Florian Klein and Dr. Matthias Zehner | AD169 strain |

**TABLE S2. UNIVARIATE ANALYSIS OF ALL IMMUNE MARKERS**

| Analyte | n | # Informative Clusters | Log OR | SE | LR test | LR P-value | FDR-adjusted P-value | Significant | Confidence Interval |
|---|---|---|---|---|---|---|---|---|---|
| **Category: Antigen ELISA** | | | | | | | | | |
| gB AD-2 (AUC) | 211 | 69 | -0.2391 | 0.1750 | 2.0062 | 0.157 | 0.313 | No | -0.5821; 0.104 |
| gB AD-6 (AUC) | 215 | 71 | -0.0457 | 0.1477 | 0.0987 | 0.753 | 0.753 | No | -0.3351; 0.2438 |
| **Category: Post Hoc ELISA** | | | | | | | | | |
| gB AD-4+5, IgG (AUC) | 210 | 69 | -1.1723 | 0.3165 | 23.4879 | <.001 | | | -1.793; -0.5519 |
| UL16, IgG (AUC) | 209 | 67 | -0.9452 | 0.2837 | 19.6071 | <.001 | | | -1.501; -0.3892 |
| prefusion-like gB, IgG (AUC) | 211 | 67 | -0.2530 | 0.1644 | 2.4933 | 0.114 | | | -0.5753; 0.06922 |
| **Category: ADCP** | | | | | | | | | |
| ADCP, AD169r (%AF647+) | 199 | 68 | -0.2333 | 0.1600 | 2.1914 | 0.139 | 0.278 | No | -0.5469; 0.08031 |
| ADCP, Towne (%AF647+) | 214 | 71 | -0.0546 | 0.1740 | 0.0983 | 0.754 | 0.754 | No | -0.3957; 0.2864 |
| **Category: Whole-Virion ELISA** | | | | | | | | | |
| AD169r virion avidity (AI%) | 195 | 68 | -0.6175 | 0.1824 | 12.9880 | <.001 | <.001 | Yes | -0.9749; -0.26 |
| AD169r virion | 216 | 72 | 0.2183 | 0.1642 | 1.9515 | 0.162 | 0.162 | No | -0.1036; 0.5402 |

| Analyte | n | # Informative Clusters | Log OR | SE | LR test | LR P-value | FDR-adjusted P-value | Significant | Confidence Interval |
|---|---|---|---|---|---|---|---|---|---|
| binding (ED50) | | | | | | | | | |
| **Category: Neutralization** | | | | | | | | | |
| Neut, AD169r, fib (ID50) | 216 | 73 | -0.3756 | 0.1673 | 5.6520 | 0.0174 | 0.0349 | Yes | -0.7035; -0.04775 |
| Neut, AD169r, epi (ID50) | 208 | 70 | 0.3349 | 0.1973 | 3.5951 | 0.058 | 0.058 | No | -0.05184; 0.7216 |
| **Category: Binding to Cell-Associated gB** | | | | | | | | | |
| cell-assoc gB IgG (%GFP+PE+) | 215 | 72 | 0.6366 | 0.2652 | 7.9147 | 0.0049 | | | 0.1169; 1.156 |
| **Category: Inhibition of Cell-Associated Viral Spread** | | | | | | | | | |
| Inhib cell-assoc spread (%sp) | 213 | 71 | -0.0078 | 0.1597 | 0.0024 | 0.961 | | | -0.3208; 0.3051 |
| **Category: DNAemia** | | | | | | | | | |
| DNAemia (IE1 copies/mL) | 216 | 72 | 0.8647 | 0.3250 | 7.5965 | 0.00585 | | | 0.2278; 1.502 |
| **Category: Multiplex Binding, IgA** | | | | | | | | | |
| pp65 (MFI) | 215 | 73 | 0.5184 | 0.1841 | 9.5418 | 0.00201 | 0.0301 | Yes | 0.1576; 0.8793 |
| gH/gL (MFI) | 216 | 74 | -0.2823 | 0.1459 | 3.9755 | 0.0462 | 0.31 | No | -0.5682; 0.00365 |

| Analyte | n | # Informative Clusters | Log OR | SE | LR test | LR P-value | FDR-adjusted P-value | Significant | Confidence Interval |
|---|---|---|---|---|---|---|---|---|---|
| gB AD-5 (MFI) | 217 | 74 | -0.2978 | 0.1711 | 3.3338 | 0.0679 | 0.31 | No | -0.633; 0.03749 |
| gB AD-1 (MFI) | 215 | 74 | 0.2618 | 0.1547 | 3.0102 | 0.0827 | 0.31 | No | -0.04145; 0.5651 |
| gB ecto (MFI) | 216 | 74 | 0.2489 | 0.1585 | 2.5437 | 0.111 | 0.323 | No | -0.06185; 0.5596 |
| full postfus gB (MFI) | 216 | 74 | 0.2422 | 0.1613 | 2.3031 | 0.129 | 0.323 | No | -0.07399; 0.5585 |
| pp150 (MFI) | 215 | 73 | 0.1814 | 0.1473 | 1.5163 | 0.218 | 0.468 | No | -0.1072; 0.4701 |
| UL16 (MFI) | 217 | 74 | 0.1558 | 0.1461 | 1.1277 | 0.288 | 0.541 | No | -0.1306; 0.4421 |
| PC (MFI) | 216 | 74 | 0.1349 | 0.1519 | 0.7925 | 0.373 | 0.584 | No | -0.1628; 0.4326 |
| gH/gL/gO (MFI) | 216 | 74 | -0.1264 | 0.1474 | 0.7413 | 0.389 | 0.584 | No | -0.4153; 0.1624 |
| UL141 (MFI) | 216 | 73 | 0.0872 | 0.1478 | 0.3450 | 0.557 | 0.694 | No | -0.2025; 0.3769 |
| prefus-like gB (MFI) | 216 | 74 | 0.0730 | 0.1433 | 0.2588 | 0.611 | 0.694 | No | -0.208; 0.3539 |
| pp71 (MFI) | 214 | 72 | 0.0634 | 0.1434 | 0.1946 | 0.659 | 0.694 | No | -0.2177; 0.3445 |
| gB AD-4 (MFI) | 216 | 74 | -0.0587 | 0.1395 | 0.1767 | 0.674 | 0.694 | No | -0.3321; 0.2148 |
| gB AD-4+5 (MFI) | 216 | 74 | -0.0638 | 0.1629 | 0.1553 | 0.694 | 0.694 | No | -0.3831; 0.2554 |

**Category: Multiplex Binding, IgG**

| Analyte | n | # Informative Clusters | Log OR | SE | LR test | LR P-value | FDR-adjusted P-value | Significant | Confidence Interval |
|---|---|---|---|---|---|---|---|---|---|
| gB AD-1 (MFI) | 217 | 74 | 0.9940 | 0.1984 | 33.3918 | <.001 | <.001 | Yes | 0.6051; 1.383 |
| pp150 (MFI) | 219 | 74 | 0.7510 | 0.1943 | 20.1497 | <.001 | <.001 | Yes | 0.3702; 1.132 |
| gB AD-4+5 (MFI) | 217 | 73 | -0.6624 | 0.1976 | 12.7486 | <.001 | 0.00183 | Yes | -1.05; -0.2752 |
| gB AD-5 (MFI) | 218 | 74 | -0.6036 | 0.1917 | 12.2846 | <.001 | 0.00183 | Yes | -0.9794; -0.2279 |
| UL16 (MFI) | 218 | 74 | -0.4866 | 0.1632 | 10.2080 | 0.0014 | 0.00447 | Yes | -0.8064; -0.1667 |
| gH/gL (MFI) | 218 | 74 | -0.4738 | 0.1724 | 8.5023 | 0.00355 | 0.00946 | Yes | -0.8117; -0.136 |
| UL141 (MFI) | 218 | 74 | -0.3569 | 0.1816 | 4.0445 | 0.0443 | 0.0945 | Yes | -0.7129; -0.0008837 |
| prefus-like gB (MFI) | 219 | 74 | -0.2998 | 0.1539 | 3.9363 | 0.0473 | 0.0945 | Yes | -0.6015; 0.001877 |
| gB AD-4 (MFI) | 219 | 74 | -0.2516 | 0.1447 | 3.0599 | 0.0802 | 0.143 | No | -0.5353; 0.03198 |
| pp65 (MFI) | 218 | 74 | 0.2130 | 0.1568 | 1.8883 | 0.169 | 0.271 | No | -0.09435; 0.5204 |
| pp71 (MFI) | 219 | 74 | -0.1301 | 0.1540 | 0.7224 | 0.395 | 0.575 | No | -0.432; 0.1718 |
| gH/UL116 (MFI) | 219 | 74 | -0.1042 | 0.1503 | 0.4787 | 0.489 | 0.637 | No | -0.3987; 0.1904 |
| PC (MFI) | 218 | 74 | 0.1232 | 0.1962 | 0.4193 | 0.517 | 0.637 | No | -0.2614; 0.5078 |

| Analyte | n | # Informative Clusters | Log OR | SE | LR test | LR P-value | FDR-adjusted P-value | Significant | Confidence Interval |
|---|---|---|---|---|---|---|---|---|---|
| gH/gL/gO (MFI) | 216 | 72 | -0.0410 | 0.1823 | 0.0503 | 0.822 | 0.879 | No | -0.3984; 0.3163 |
| full postfus gB (MFI) | 217 | 73 | -0.0432 | 0.1945 | 0.0492 | 0.824 | 0.879 | No | -0.4244; 0.3379 |
| gB ecto (MFI) | 218 | 74 | -0.0119 | 0.1888 | 0.0039 | 0.95 | 0.95 | No | -0.3819; 0.3582 |
| **Category: Multiplex Binding, IgM** | | | | | | | | | |
| gB ecto (MFI) | 214 | 71 | 0.8962 | 0.1797 | 33.9965 | <.001 | <.001 | Yes | 0.5441; 1.248 |
| PC (MFI) | 214 | 71 | 0.8345 | 0.1804 | 28.9969 | <.001 | <.001 | Yes | 0.481; 1.188 |
| full postfus gB (MFI) | 214 | 71 | 0.4844 | 0.1429 | 12.4150 | <.001 | 0.00207 | Yes | 0.2044; 0.7645 |
| gB AD-1 (MFI) | 200 | 67 | 0.5743 | 0.1836 | 11.9289 | <.001 | 0.00207 | Yes | 0.2145; 0.9341 |
| pp150 (MFI) | 200 | 67 | 0.5750 | 0.1860 | 11.4611 | <.001 | 0.00213 | Yes | 0.2105; 0.9396 |
| pp71 (MFI) | 200 | 67 | 0.4946 | 0.1768 | 8.8630 | 0.00291 | 0.00728 | Yes | 0.1481; 0.8411 |
| UL16 (MFI) | 202 | 67 | 0.5210 | 0.2341 | 7.0074 | 0.00812 | 0.0174 | Yes | 0.06221; 0.9798 |
| gB AD-4+5 (MFI) | 202 | 67 | 0.3477 | 0.1521 | 5.4799 | 0.0192 | 0.0361 | Yes | 0.04954; 0.6458 |
| gB AD-4 (MFI) | 202 | 67 | 0.3004 | 0.1577 | 3.8780 | 0.0489 | 0.0761 | Yes | -0.008824; 0.6095 |
| prefus-like gB (MFI) | 202 | 67 | 0.3081 | 0.1604 | 3.8180 | 0.0507 | 0.0761 | No | -0.006246; 0.6224 |

| Analyte | n | # Informative Clusters | Log OR | SE | LR test | LR P-value | FDR-adjusted P-value | Significant | Confidence Interval |
|---|---|---|---|---|---|---|---|---|---|
| gH/gL/gO (MFI) | 214 | 71 | 0.2682 | 0.1423 | 3.6520 | 0.056 | 0.0764 | No | -0.01075; 0.5471 |
| gB AD-5 (MFI) | 202 | 67 | 0.1781 | 0.1542 | 1.3527 | 0.245 | 0.306 | No | -0.1241; 0.4803 |
| gH/gL (MFI) | 202 | 67 | 0.1457 | 0.1445 | 0.9995 | 0.317 | 0.354 | No | -0.1375; 0.429 |
| pp65 (MFI) | 200 | 67 | 0.1438 | 0.1473 | 0.9482 | 0.33 | 0.354 | No | -0.1449; 0.4325 |
| UL141 (MFI) | 199 | 66 | 0.1309 | 0.1604 | 0.6548 | 0.418 | 0.418 | No | -0.1835; 0.4453 |

**TABLE S3. UNIVARIATE ANALYSIS OF ALL SIGNIFICANT IMMUNE MARKERS CONDITIONED ON THE PRESENCE OF DNAEMIA**

| Analyte | # Informative Clusters | Adjusted Log OR | Non Adjusted Log OR | Adjusted LR P-value | Non Adjusted LR P-value | Adjusted Confidence Interval | Non Adjusted Confidence Interval |
|---|---|---|---|---|---|---|---|
| **Category: Whole-Virion ELISA** | | | | | | | |
| AD169r virion avidity (AI%) | 66 | -0.4802 | -0.5624 | 0.00967 | 0.00156 | -0.8566; -0.1037 | -0.9285; -0.1962 |
| **Category: Neutralization** | | | | | | | |
| Neut, AD169r, fib (ID50) | 71 | -0.4196 | -0.3849 | 0.0133 | 0.0189 | -0.7764; -0.0629 | -0.7274; -0.04244 |
| **Category: Binding to Cell-Associated gB** | | | | | | | |
| cell-assoc gB IgG (%GFP+PE+) | 70 | 0.5728 | 0.6371 | 0.0198 | 0.00567 | 0.01831; 1.127 | 0.1057; 1.169 |
| **Category: Post Hoc ELISA** | | | | | | | |
| gB AD-4+5, IgG (AUC) | 67 | -1.1084 | -1.1169 | <.001 | <.001 | -1.726; -0.4911 | -1.726; -0.5076 |
| UL16, IgG (AUC) | 66 | -0.8912 | -0.9525 | <.001 | <.001 | -1.456; -0.3262 | -1.512; -0.3934 |
| **Category: Multiplex Binding, IgA** | | | | | | | |
| pp65 (MFI) | 71 | 0.4649 | 0.4876 | 0.00998 | 0.00489 | 0.08107; 0.8487 | 0.119; 0.8563 |
| **Category: Multiplex Binding, IgG** | | | | | | | |
| gB AD-1 (MFI) | 72 | 0.9498 | 0.9882 | <.001 | <.001 | 0.5516; 1.348 | 0.5946; 1.382 |

| Analyte | # Informative Clusters | Adjusted Log OR | Non Adjusted Log OR | Adjusted LR P-value | Non Adjusted LR P-value | Adjusted Confidence Interval | Non Adjusted Confidence Interval |
|---|---|---|---|---|---|---|---|
| pp150 (MFI) | 72 | 0.6549 | 0.7063 | <.001 | <.001 | 0.2657; 1.044 | 0.3272; 1.085 |
| gB AD-4+5 (MFI) | 71 | -0.6641 | -0.6762 | <.001 | <.001 | -1.061; -0.2676 | -1.07; -0.2824 |
| gB AD-5 (MFI) | 72 | -0.5391 | -0.5758 | 0.00285 | <.001 | -0.927; -0.1513 | -0.9537; -0.1979 |
| gH/gL (MFI) | 72 | -0.4945 | -0.4589 | 0.00364 | 0.00548 | -0.842; -0.147 | -0.8013; -0.1165 |
| UL16 (MFI) | 72 | -0.4724 | -0.5224 | 0.00389 | <.001 | -0.8136; -0.1312 | -0.8547; -0.1901 |
| UL141 (MFI) | 72 | -0.3620 | -0.3412 | 0.0566 | 0.0565 | -0.7427; 0.01868 | -0.6997; 0.01737 |
| prefus-like gB (MFI) | 72 | -0.2864 | -0.2777 | 0.0708 | 0.0694 | -0.5953; 0.02258 | -0.5814; 0.0261 |
| **Category: Multiplex Binding, IgM** | | | | | | | |
| gB ecto (MFI) | 69 | 0.8505 | 0.8648 | <.001 | <.001 | 0.4953; 1.206 | 0.5132; 1.216 |
| PC (MFI) | 69 | 0.7971 | 0.8002 | <.001 | <.001 | 0.4383; 1.156 | 0.4491; 1.151 |
| full postfus gB (MFI) | 69 | 0.5073 | 0.4727 | <.001 | <.001 | 0.2095; 0.8051 | 0.1901; 0.7554 |
| pp150 (MFI) | 65 | 0.5259 | 0.5460 | 0.00226 | 0.00141 | 0.1599; 0.8919 | 0.1808; 0.9111 |
| gB AD-1 (MFI) | 65 | 0.5325 | 0.5619 | 0.00261 | 0.00106 | 0.1603; 0.9047 | 0.1965; 0.9272 |

| Analyte | # Informative Clusters | Adjusted Log OR | Non Adjusted Log OR | Adjusted LR P-value | Non Adjusted LR P-value | Adjusted Confidence Interval | Non Adjusted Confidence Interval |
|---|---|---|---|---|---|---|---|
| pp71 (MFI) | 65 | 0.5356 | 0.5093 | 0.00297 | 0.00383 | 0.1607; 0.9105 | 0.144; 0.8747 |
| UL16 (MFI) | 65 | 0.6068 | 0.5210 | 0.00424 | 0.00812 | 0.1117; 1.102 | 0.06221; 0.9798 |
| gB AD-4+5 (MFI) | 65 | 0.2856 | 0.3371 | 0.0617 | 0.0235 | -0.01937; 0.5907 | 0.03921; 0.6351 |
| gB AD-4 (MFI) | 65 | 0.2443 | 0.2285 | 0.163 | 0.172 | -0.1033; 0.5918 | -0.1023; 0.5593 |

**TABLE S4. ANALYSIS OF IMPACT OF DNAEMIA MAGNITUDE ON TRANMISSION ODDS**

| Analyte | n | Informative Clusters | LogOR | SE | Wald P-value | Confidence Interval |
|---|---|---|---|---|---|---|
| DNAemia (IE1 copies/mL) | 216 | 72 | 3.0122 | 1.4514 | 0.038 | 0.1674; 5.857 |
| log10 VL Magnitude | 216 | 72 | -0.3109 | 0.2047 | 0.129 | -0.7121; 0.09037 |

**TABLE S5. COMPARISON OF IMMUNE MARKERS IN PARTICIPANTS WITH AND WITHOUT CONCURRENT DNAEMIA**

| Immune Marker | P-value, Wilcoxon Rank Sum Test |
|---|---|
| AD169r virion avidity, IgG (AI%) | 0.3088 |
| AD169r virion binding, IgG (ED50) | 0.2102 |
| ADCP, AD169r (%AF647+) | 0.9954 |
| ADCP, Towne (%AF647+) | 0.7187 |
| Inhibition of cell-associated viral spread (% spread) | 0.0181 |
| Neutralization, AD169r, epithelial cells (ID50) | 0.0212 |
| Neutralization, AD169r, fibroblasts (ID50) | 0.4601 |
| UL141 binding, IgA (MFI) | 0.4240 |
| UL141 binding, IgG (MFI) | 0.9124 |
| UL141 binding, IgM (MFI) | 0.0395 |
| UL16 binding, IgA (MFI) | 0.9159 |
| UL16 binding, IgG (AUC) | 0.0396 |
| UL16 binding, IgG (MFI) | 0.0269 |
| UL16 binding, IgM (MFI) | 0.0555 |
| binding to cell-associated gB (%GFP+PE+) | 0.0813 |
| full-length postfusion gB binding, IgA (MFI) | 0.0225 |
| full-length postfusion gB binding, IgG (MFI) | 0.7352 |
| full-length postfusion gB binding, IgM (MFI) | 0.9763 |
| gB AD-1 binding, IgA (MFI) | 0.4114 |
| gB AD-1 binding, IgG (MFI) | 0.3429 |
| gB AD-1 binding, IgM (MFI) | 0.7785 |
| gB AD-2 binding, IgG (AUC) | 0.5666 |
| gB AD-4 binding, IgA (MFI) | 0.5169 |
| gB AD-4 binding, IgG (MFI) | 0.8311 |
| gB AD-4 binding, IgM (MFI) | 0.6546 |
| gB AD-4+5 binding, IgA (MFI) | 0.0047 |
| gB AD-4+5 binding, IgG (AUC) | 0.6951 |
| gB AD-4+5 binding, IgG (MFI) | 0.1891 |

| Immune Marker | P-value, Wilcoxon Rank Sum Test |
|---|---:|
| gB AD-4+5 binding, IgM (MFI) | 0.6074 |
| gB AD-5 binding, IgA (MFI) | 0.3320 |
| gB AD-5 binding, IgG (MFI) | 0.2305 |
| gB AD-5 binding, IgM (MFI) | 1.0000 |
| gB AD-6 binding, IgG (AUC) | 0.7730 |
| gB ectodomain binding, IgA (MFI) | 0.2096 |
| gB ectodomain binding, IgG (MFI) | 0.2292 |
| gB ectodomain binding, IgM (MFI) | 0.2125 |
| gH/UL116 binding, IgG (MFI) | 0.2188 |
| gH/gL binding, IgA (MFI) | 0.5935 |
| gH/gL binding, IgG (MFI) | 0.5002 |
| gH/gL binding, IgM (MFI) | 0.6932 |
| gH/gL/gO binding, IgA (MFI) | 0.0156 |
| gH/gL/gO binding, IgG (MFI) | 0.0598 |
| gH/gL/gO binding, IgM (MFI) | 0.3097 |
| pentameric complex binding, IgA (MFI) | 0.0286 |
| pentameric complex binding, IgG (MFI) | 0.0111 |
| pentameric complex binding, IgM (MFI) | 0.2130 |
| pp150 binding, IgA (MFI) | 0.3439 |
| pp150 binding, IgG (MFI) | 0.0054 |
| pp150 binding, IgM (MFI) | 0.7417 |
| pp65 binding, IgA (MFI) | 0.6899 |
| pp65 binding, IgG (MFI) | 0.6363 |
| pp65 binding, IgM (MFI) | 0.1611 |
| pp71 binding, IgA (MFI) | 0.3089 |
| pp71 binding, IgG (MFI) | 0.2340 |
| pp71 binding, IgM (MFI) | 0.6807 |
| prefusion gB binding, IgA (MFI) | 0.8071 |
| prefusion-like gB binding, IgG (AUC) | 0.3424 |
| prefusion-like gB binding, IgG (MFI) | 0.5642 |

| Immune Marker | P-value, Wilcoxon Rank Sum Test |
| --- | --- |
| prefusion-like gB binding, IgM (MFI) | 0.1809 |

### TABLE S6. SPEARMAN CORRELATION FOR ALL IMMUNE MARKERS

Attached as a .csv file.

**TABLE S7. OVERALL REPRESENTATIVENESS OF THE TRIAL**

| Category | Description |
|---|---|
| Disease, problem, or condition under investigation | Primary human cytomegalovirus (HCMV) infection prior to 24 weeks gestation; vertical transmission of HCMV |
| Special considerations related to | |
| Sex and gender | Pregnant individuals; little data regarding the impact of fetal sex on transmission risk with one study suggesting fetal sex is not associated with transmission risk[17]. |
| Age | Mean maternal age at birth in the United States during the trial period (2012-2018) was 28.5 years[18]. Younger maternal age is associated with increased risk of congenital CMV transmission[19]. |
| Race or ethnic group | Congenital CMV rates are higher in black infants in the United States[20] |
| Geography | HCMV seroprevalence in the United States is approximately 40-60% for women of reproductive age[21]. Global seroprevalence rates vary and are >95% in some regions[22]. |
| Other considerations | Exposure to young children is known risk factor for congenital CMV as children are a primary source of HCMV transmission[23]. |
| Overall representativeness of this trial | While the incidence of congenital CMV in the United States has been estimated, the incidence of primary infection in the United States is difficult to estimate due a lack of routine maternal screening. This study, which screened over 200,000 pregnancies in the United States, provided the most comprehensive assessment of vertical transmission following primary HCMV in pregnancy in the United States to date[1,24]. While this trial likely accurately reflects the population of women with primary infection during pregnancy in the United States, global seroprevalence rates vary significantly and this may not be representative of women with primary infection in other countries.<br><br>Trial participants were all female sex since the trial was conducted during pregnancy; fetal sex was not reported in this trial. Participants in our case-control cohort had mean ages of 27.2 years (Non-transmitters) and 26.4 years (Transmitters). While participants in the trial were younger than the national mean age, younger maternal age is associated with increased incidence of cCMV. Participants were enrolled at 16 sites distributed throughout the United States. Participants were matched on the following demographic features: maternal age (within 10 years), maternal race (White, Black, Mixed Race/Other/Not reported), maternal ethnicity (Hispanic/Latino, Non-Hispanic/Latino), and parity (multiparous, nulliparous). |

**REFERENCES**

1. Hughes BL, Clifton RG, Rouse DJ, et al. A Trial of Hyperimmune Globulin to Prevent Congenital Cytomegalovirus Infection. N Engl J Med 2021;385(5):436–444. DOI: 10.1056/NEJMoa1913569.
2. Bialas KM, Westreich D, Cisneros de la Rosa E, et al. Maternal Antibody Responses and Nonprimary Congenital Cytomegalovirus Infection of HIV-1-Exposed Infants. J Infect Dis 2016;214(12):1916–1923. DOI: 10.1093/infdis/jiw487.
3. Nelson CS, Huffman T, Jenks JA, et al. HCMV glycoprotein B subunit vaccine efficacy mediated by nonneutralizing antibody effector functions. Proceedings of the National Academy of Sciences 2018;115(24):6267–6272. DOI: 10.1073/pnas.1800177115.
4. Nelson CS, Jenks JA, Pardi N, et al. Human Cytomegalovirus Glycoprotein B Nucleoside-Modified mRNA Vaccine Elicits Antibody Responses with Greater Durability and Breadth than MF59-Adjuvanted gB Protein Immunization. J Virol 2020;94(9). DOI: 10.1128/JVI.00186-20.
5. Semmes EC, Miller IG, Wimberly CE, et al. Maternal Fc-mediated non-neutralizing antibody responses correlate with protection against congenital human cytomegalovirus infection. J Clin Invest 2022;132(16). DOI: 10.1172/JCI156827.
6. Hu X, Karthigeyan KP, Herbek S, et al. Human Cytomegalovirus mRNA-1647 Vaccine Candidate Elicits Potent and Broad Neutralization and Higher Antibody-Dependent Cellular Cytotoxicity Responses Than the gB/MF59 Vaccine. J Infect Dis 2024;230(2):455–466. DOI: 10.1093/infdis/jiad593.
7. Karthigeyan KP, Connors M, Binuya CR, et al. A human cytomegalovirus prefusion-like glycoprotein B subunit vaccine elicits humoral immunity similar to that of postfusion gB in mice. J Virol 2025:e0217824. DOI: 10.1128/jvi.02178-24.
8. Connors MR, Karthigeyan KP, Fuller AS, et al. Specificity and functional humoral immune responses induced by the VBI-1501A eVLP HCMV gB vaccine compared to the gB/MF59 vaccine. Hum Vaccin Immunother 2025;21(1):2564555. DOI: 10.1080/21645515.2025.2564555.
9. Jenks JA, Nelson CS, Roark HK, et al. Antibody binding to native cytomegalovirus glycoprotein B predicts efficacy of the gB/MF59 vaccine in humans. Sci Transl Med 2020;12(568):eabb3611. DOI: 10.1126/scitranslmed.abb3611.
10. Rouse DJ, Fette LM, Hughes BL, et al. Noninvasive Prediction of Congenital Cytomegalovirus Infection After Maternal Primary Infection. Obstet Gynecol 2022;139(3):400–406. DOI: 10.1097/AOG.0000000000004691.
11. Sekhon JS. Multivariate and Propensity Score Matching Software with Automated Balance Optimization: The Matching Package for R. J Stat Softw 2011;42(7):1–52. (In English) (<Go to ISI>://WOS:000292097200001).
12. Therneau T. A package for survival analysis in R. (https://cran.r-project.org/web/packages/survival/index.html).
13. Gu ZG. Complex heatmap visualization. Imeta 2022;1(3) (In English). DOI: ARTN e43 10.1002/imt2.43.
14. Hothon T, Winell H, Hornik K, van de Wiel MA, Zeileis A. Conditional Inference Procedures in a Permutation Test Framework.
15. Reid S, Tibshirani R. Regularization Paths for Conditional Logistic Regression: The clogitL1 Package. J Stat Softw 2014;58(12):1–23. (In English) (<Go to ISI>://WOS:000341642900001).
16. van Buuren S, Groothuis-Oudshoorn K. mice: Multivariate Imputation by Chained Equations in R. J Stat Softw 2011;45(3):1–67. (In English) (<Go to ISI>://WOS:000298032500001).
17. Garozzo MT, Pecorino B, Poli G, et al. Maternal transmission, neonatal outcomes, and predictors of adverse effects in congenital cytomegalovirus infection. Pediatr Neonatol 2026;67(5):523–531. DOI: 10.1016/j.pedneo.2025.08.016.
18. Martin JA, Hamilton BE, Osterman MJK, Driscoll A. Births: Final Data for 2018. National Vital Statistics Reports. Hyattsville, MD: 2019.
19. Fowler KB, Stagno S, Pass RF. Maternal age and congenital cytomegalovirus infection: screening of two diverse newborn populations, 1980-1990. J Infect Dis 1993;168(3):552–6. DOI: 10.1093/infdis/168.3.552.
20. Fowler KB, Ross SA, Shimamura M, et al. Racial and Ethnic Differences in the Prevalence of Congenital Cytomegalovirus Infection. J Pediatr 2018;200:196–201 e1. DOI: 10.1016/j.jpeds.2018.04.043.

21. Dana Flanders W, Lally C, Dilley A, Diaz-Decaro J. Estimated cytomegalovirus seroprevalence in the general population of the United States and Canada. J Med Virol 2024;96(3):e29525. DOI: 10.1002/jmv.29525.
22. Boppana SB, van Boven M, Britt WJ, et al. Vaccine value profile for cytomegalovirus. Vaccine 2023;41 Suppl 2:S53–S75. DOI: 10.1016/j.vaccine.2023.06.020.
23. CDC. Cytomegalovirus (CMV) and Congenital CMV Infection. (https://www.cdc.gov/cytomegalovirus/about/index.html).
24. Hughes BL, MacPherson C, Rouse DJ, et al. Maternal cytomegalovirus serology results by geographic region in a large multicenter United States study. J Infect Dis 2026. DOI: 10.1093/infdis/jiag224.
